# Data Analysis and Forecasting of COVID-19 Pandemic in Kuwait

**DOI:** 10.1101/2021.07.24.21261059

**Authors:** Kayode Oshinubi, Fahimah Al-Awadhi, Mustapha Rachdi, Jacques Demongeot

## Abstract

The first COVID 19 case of Kuwait was announced on 24th February, 2020 and the daily new cases increases exponentially since then until May, 2020 when the first wave started to decline. The same exponential dynamics has been observed between January and March, 2021. The forecast of new cases and death recorded daily is crucial so that health experts and citizens can be guided in order to avoid escalation of the pandemic. We propose a deterministic method to predict the basic reproduction number *R*_*o*_ of first and second wave of COVID-19 cases in Kuwait and also to forecast the daily new cases and death of the pandemic in the country. Forecasting has been done using ARIMA model, Exponential smoothing model, Holts’ method, Prophet forecasting model and machine learning models like log-linear, polynomial and support vector regressions. The results presented aligned with other methods used to predict *R*_*o*_ in first and second waves and the forecasting clearly shows the trend of the pandemic in Kuwait. The deterministic prediction of *R*_*o*_ is a good forecasting tool available during the exponential phase of the contagion, which shows an increasing trend during the beginning of the first and second waves of the pandemic in Kuwait.

## 1. Introduction

In January, 2020 WHO declared the coronavirus is responsible for respiratory illness in Wuhan China and the first case in Kuwait was reported on 24th February, 2020. Since this emergence of COVID-19 pandemic in Kuwait, the country has experienced a high stationary number of new cases since the end of the first wave in May, 2020 until a slight decline in December, 2020. After, the new cases in the second wave are on exponential increase since January, 2021 until March, 2020 followed by a stationary phase analogue to that of 2020 but with a higher average number of daily new cases.

The COVID-19 pandemic has resulted in a lot of fatality across the globe, but the tenth of April 2021, the statistics by worldometer® (Worldometer 2021) indicated that Kuwait cumulated number of cases is 244,325, with a number of deaths equal to 1,393 (323 per million), corresponding to a fatality rate of 0.58%, placing Kuwait in 83rd place (in the sense of decreasing fatality rate) of developed countries. Recovery rate is 93.57%, critical cases treated in ICU are 224 − 0.09% of total cases, daily cases receiving treatment is 14, 305 − 5.85% of total cases and cumulated confirmed cases proportion equals 56,568 per million.

In recent time, there has been a lot of research on the COVID-19 pandemic in different fields ranging from statistics, epidemiology, mathematics, biology, medicine, etc. and these fields have looked at various aspects of COVID-19 pandemic modelling in the areas reported and unreported cases, prediction of *R*_*o*_, lockdown and more recently introduction of vaccines. For example, in (Demongeot *et al*. 2020) authors worked on the spread parameters of the new COVID-19 cases dynamics and conclude on how temperature indicates the cases in 21 countries and French administrative regions. They proposed ARIMA model to analyse incidence pattern and estimate short-term forecasts for retro-predicting the first wave of COVID-19 outbreak. Inverted covariate effects for first versus mutated second wave COVID-19 and how confinements hasten viral evolution toward greater contagiousness were investigated in (Seligmann *et al*. 2020), and a new method for calculating the daily reproduction number during the contagiousness period of an individual has been also recently proposed (Demongeot *et al*. 2021).

We propose in the present paper some data analysis tools for Kuwait in terms of daily new cases and deaths in the country. In Section 2 we describe the methods used in this paper for processing data and present the deterministic modelling on the daily new cases observed in order to predict *R*_*o*_. In Section 3, we give results and visualisation of the machine learning tools applied on the pattern of the daily new cases and deaths in Kuwait. Eventually, in Section 4, we conclude by giving some perspectives about the generalisation of the present work to other countries.

## 2. Materials and Methods

We use various time series forecasting techniques like ARIMA and machine learning models like loglinear, polynomial and support vector regressions deployed to study the daily new cases dynamics and also the daily number of deaths recorded and forecasted in the near future. We also use exponential smoothing method, Holt’s method and Prophet forecast modelling to validate our results. We took 100 days both in the first wave and second wave depending on the available data. First wave new cases were considered from 25/02/2020 to 03/06/2020 while second wave new cases were considered from 15/10/2020 to 22/01/2021. For the first wave, we used daily deaths data from 04/04/2020 to 12/07/2020, while for the second wave we used those from 15/10/2020 to 22/01/2021. We also calculated the slopes from the log-linear regression analysis using exponential model *y* = *ae*^*bx*^, where *y* is the daily number of new cases, *x* the number of days, *b* the slope and *loga* a constant in the log format *logy* = *loga* + *bx*. We also calculated the initial negative autocorrelation slope of the epidemic spread averaged on six days. A deterministic model was used to predict *R*_*o*_ of both the first and second wave of the pandemic in Kuwait. Simulation, data visualisation and computation were done in a Python environment.

### 2.1 Methods

#### 2.1.1 Time series modelling

Time series modelling has been introduced by N. Wiener for prediction and forecasting (Wiener 1949). Its parametric approach assumes that after subtracting the trend (increasing or decreasing) of Covid-19 new cases dynamics, we get an underlying stationary stochastic process *N* (*t*), which can be described by a small number of parameters using the autoregressive ARIMA model:

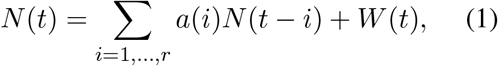

where *W* is a random residue, whose variance is to minimise. The autocorrelation curve is obtained by calculating the correlation *A*(*k*) between *N* (*t*) and the *N* (*t − k*)’s (*t* belonging to a moving time window) by using the formula:

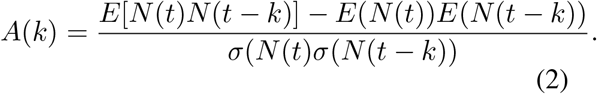

where *E* denotes the expectation and *σ* the standard deviation. The autocorrelation function *A* allows examining the serial dependence of the *N* (*t*)’s.

#### 2.1.2 Linear, Polynomial and Support Vector Regression

Log-linear regression model use some historic data between variables *X* and *Y* depending on *X*, and consider a linear relationship between *logY* and *X*, while polynomial regression models use a similar approach but the dependent variable *Y* is modelled as a polynomial of degree *n* in *X*. Support vector regression is a supervised machine learning model which draws a hyperplane relating the data points and creates a boundary of possible data points (high and low) in future. Support vector regression traditionally has huge forecasting abilities.

#### 2.1.3 The Prophet Forecasting Model, Exponential smoothing model and Holts’ model

The Prophet forecasting modelling uses a decomposable time series model with three main model components which are trend, seasonality and holidays. It is described by the model equation below:

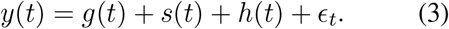

where *g*(*t*) is a piecewise linear or logistic growth curve for modelling non-periodic changes in time series, *s*(*t*) represent periodic changes like weekly, yearly or seasonal, *h*(*t*) is effects of holidays with irregular schedules and *E*_*t*_. is the error term which account for any unusual changes or noise not accommodated by the model (Rafferty 2021). Exponential smoothing methods use weighted averages of past observations to forecast new values. It combines error, trend and seasonal components in a smoothing calculation. Each term can be combined either additively, multiplicatively or be left out of the model. Holts’ winter method is also called triple exponential smoothing and it is used in order to apply exponential smoothing to the seasonal components in addition to level and trend. It is a method that takes into account both seasonality and trend when forecasting. Holt’s linear trend method takes into account the trend of a given data set. The method map the trend accurately without any assumptions (Chatfield 2000).

### 2.2 Deterministic modelling

According to (Demongeot *et al*. 2021), let *R*_*o*_ denotes the average transmission rate (or basic reproduction number) among the Kuwait population. We can estimate the distribution *V* (whose coefficients are denoted *V*_*j*_ = *R*_*j*_*/R*_*o*_) of the daily reproduction numbers *R*_*j*_ along the contagious period of an individual, by remarking that the number *X*_*j*_ of new infectious cases at day *j*, equal to *X*_*j*_ = *I*(*j*)–*I*(*j−* 1), where *I*(*j*) is the cumulated number of infectious at day *j*, verifies the discrete convolution equation:

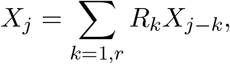

giving in continuous time:

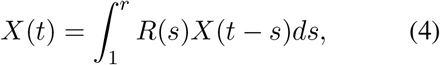

where *r* is the duration of the contagion period, estimated by 1*/*(*k* + *µ*),where *k* is the recovering rate and *µ* the death rate in SIR equations:

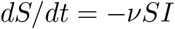

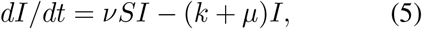

where *S* and *I* are respectively the size of the susceptible and infectious populations.

If *r* and *S* can be considered as constant during the first exponential phase of the pandemic, we can also assume that the distribution *V* is constant and then, *V* can be estimated by solving the linear system:

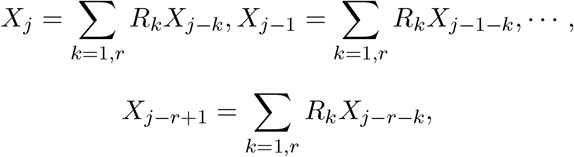

which can be written as *X* = *MR*, hence giving *R* = *M* ^−1^*X*, where:

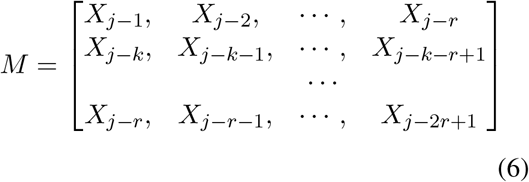

and equation (6) can be solved numerically, if the pandemic is observed during a time greater than 1*/*(*k* + *µ*). Then, the entropy of *V* = *R/R*_*o*_ can be calculated, as the Kolmogorov-Sinaï entropy of the Markovian Delbrück scheme ruling the 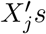 and giving new parameters for characterising pandemic dynamics, namely for quantifying its robustness and stability ((Rhodes & Demetrius 2010), (Demongeot & Demetrius 2015)).

If there are negative *V*_*j*_’s, it is still possible to define an index of proximity to uniformity of *V* by considering the entropy of the distribution *W* defined by *W*_*j*_ = [(*V*_*j*_ −*min*{*V*_*k*_ *≤* 0})*/* ∑_*i*=1,*r*_ *W*_*i*_].

#### 2.2.1 Start of the pandemic in Kuwait

If we use the daily new infectious cases given in worldometer at the start of the pandemic, we can calculate *M* ^−1^ for the period from February 25 to March 1, by choosing 3 days for the duration of the infectiousness period and the following raw data for the new infected cases are *X*_1_ = 6 the 25th February, *X*_2_ = 15 the 26th, *X*_3_ = 17 the 27th, *X*_4_ = 2 the 28th, *X*_5_ = 0 the 29th and *X*_6_ = 1 the 1st March, hence giving for the matrix M:

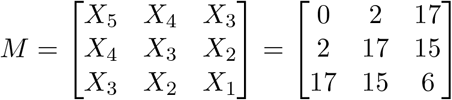

Then, we have:

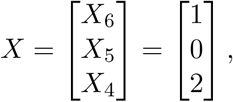

Because

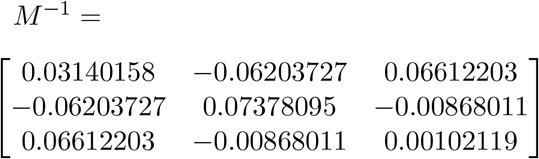

we get *R* = *M* ^−1^*X*, where:

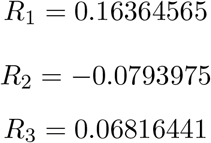

Then, we can give a graphical representation of the *R*_*j*_’s (Figure 1) and the average transmission rate *R*_*o*_ ≈ 0.2, value close to that calculated directly, with a maximal daily reproduction rate the first day of the infectiousness period. Because of the negativity of *R*_2_, we have to calculate the entropy *H* of the distribution *W* = [(*R*_1_*−R*_2_)*/*(*R*_1_−2*R*_2_ + *R*_3_), 0, (*R*_3_*− R*_2_)*/*(*R*_1_ − 2*R*_2_ + *R*_3_)] = (0.622, 0, 0.378), which equals 0.663.

**Fig. 1.**
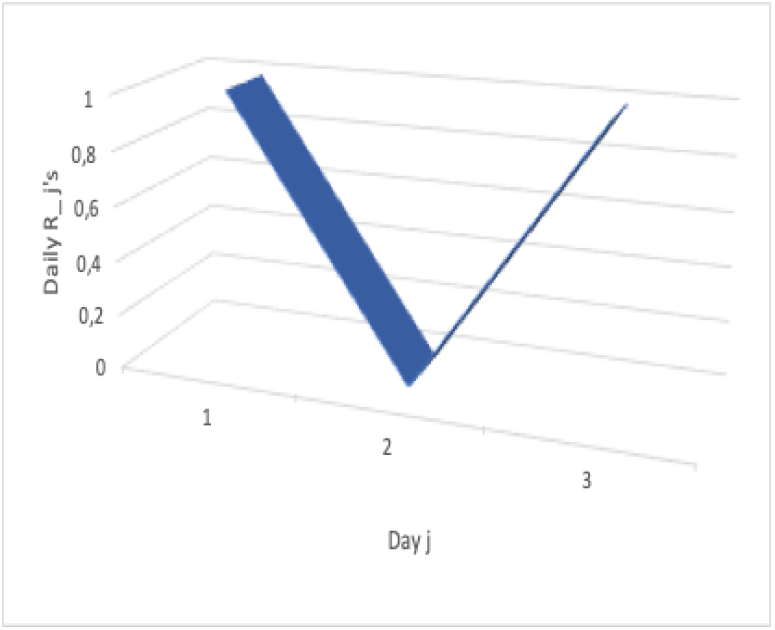
V-shape of the evolution of the daily *R*_*j*_’s along the infectious 3-day period of an individual, calculated for the period from February 25 to March 1 2020.

#### 2.2.2 First exponential phase of the pandemic in Kuwait

If we use the daily new infectious cases given in (Worldometer 2021) during the exponential phase of the first wave, we can calculate *M* ^−1^ for the period from October 20 to October 25, by choosing 3 days for the duration of the infectiousness period and the following raw data for the new infected cases are *X*_1_ = 886 the 20th October, *X*_2_ = 813 the 21th, *X*_3_ = 889 the 22th, *X*_4_ = 812 the 23th, *X*_5_ = 695 the 24th and *X*_6_ = 708 the 25th, hence giving for the matrix *M* :

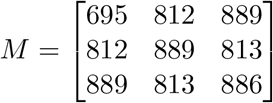

Then, we have:

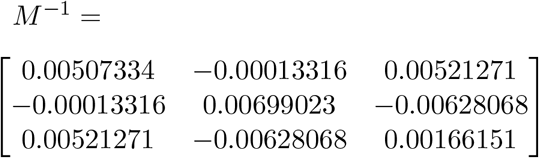

Because

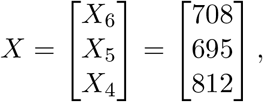

we get *R* = *M* ^−1^*X*, where:

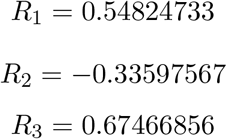

Then, we can give a graphical representation of the *R*_*j*_’s (Figure 2). The average transmission rate *R*_*o*_ verifies *R*_*o*_ ≈ 0.9, value close to that calculated directly, with a maximal daily reproduction rate the first day of the infectiousness period. Because of the negativity of*R*_2_, we have to calculate the entropy *H* of the distribution *W* = [(*R*_1_*−R*_2_)*/*(*R*_1_ − 2*R*_2_ + *R*_3_), 0, (*R*_3_*− R*_2_)*/*(*R*_1_ − 2*R*_2_ + *R*_3_)] = (0.467, 0, 0.533), which is equal to *H* = 0.691.

**Fig. 2.**
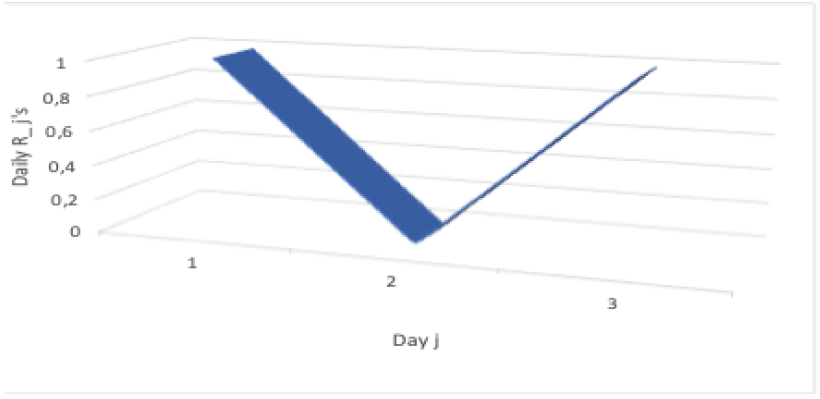
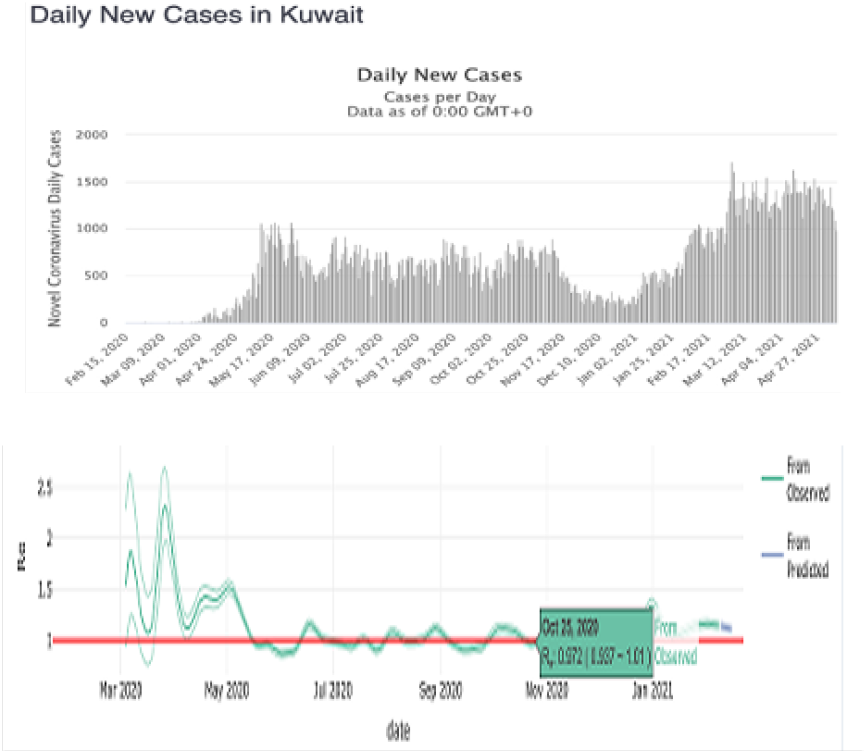
V-shape of the evolution of the daily *R*_*j*_’s along the infectious 3-day period of an individual, daily new cases in Kuwait between October 20 to October 25 2020 in worldometer and estimation of the average transmission rate *R*_*o*_ for the 20th and 25th October 2020 with its 95% confidence interval (in green).

#### 2.2.3 Second exponential phase of the pandemic in Kuwait

If we use the daily new infectious cases given in (Worldometer 2021) during the exponential phase of the second wave, we can calculate *M* ^−1^ for the period from December 30 to January 4, by choosing 3 days for the duration of the infectiousness period and the following raw data for the new infected cases are *X*_1_ = 205 the 30th December, *X*_2_ = 286 the 31th, *X*_3_ = 285 the 1st January, *X*_4_ = 205 the 2nd, *X*_5_ = 269 the 3rd and *X*_6_ = 372 the 4th, hence giving for the matrix *M* :

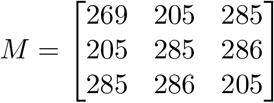

Then, we have:

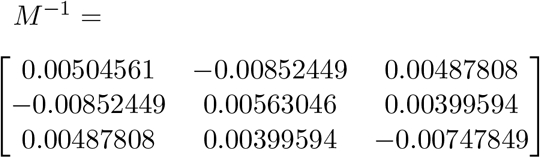

Because

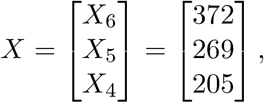

we get *R* = *M* ^−1^*X*, where:

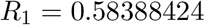

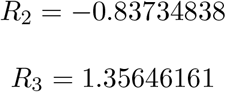

Then, we can give a graphical representation of the *R*_*j*_’s (Figure 3). The average transmission rate *R*_*o*_ equals about 1.1, value close to that calculated directly, with a maximal daily reproduction rate the first day of the infectiousness period. Because of the negativity of *R*_2_, we have to calculate the entropy *H* of the distribution *W* = [(*R*_1_*−R*_2_)*/*(*R*_1_ − 2*R*_2_ + *R*_3_), 0, (*R*_3_*− R*_2_)*/*(*R*_1_ − 2*R*_2_ + *R*_3_)] = (0.393, 0, 0.607), which is equal to *H* = 0.691, the same value than for the first wave.

**Fig. 3.**
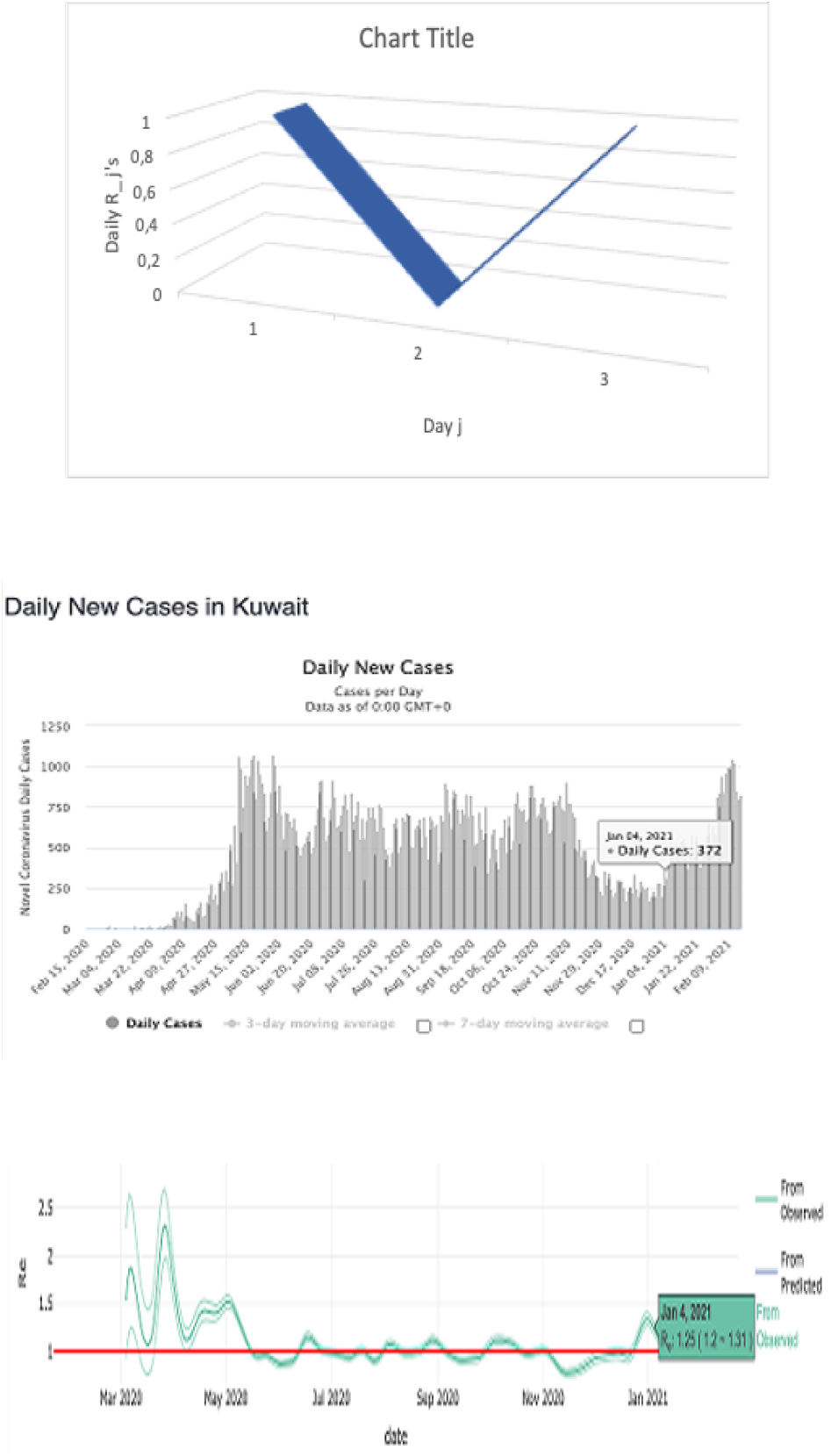
V-shape of the evolution of the daily *R*_*j*_’s along the infectious 3-day period of an individual, daily new cases in Kuwait between December 302020 and January 4 2021 in worldometer and estimation of the average transmission rate *R*_*o*_ for December 30 2020 and January 4 2021 with its 95% confidence interval (in green).

## 3. Results

### 3.1 Regression curves and ARIMA autocorrelation slopes for first wave start and second wave transition

As expected, the 100 first days of the first wave start show a positive slope (0.0687) and the 100 first days of the second wave transition show a negative slope (−0.0094).

For first wave start, we obtain the exponential model: y = 0.8e0.07x, based on the following results: Loglinear regression: slope = 0.0686852051631, intercept=0.800565672854, rvalue = 0.939152595486, pvalue = 2.079801359 e-45, stderr = 0.002591135543915, R-squared = 0.882008, RMSE = 0.711400960815. By using other types of regression, we get: Polynomial Regression: R-squared = 0.787421, RMSE = 0.586403581239, Support vector Regression: R-squared = 0.98382, RMSE = 0.263424533991. For second wave transition, the exponential model is y = 6.5e-0.009x, based on the following results: Loglinear Regression: slope = -0.0093674219195, intercept = 6.5198283885, rvalue = -0.57106504118, pvalue = 5.50386624132 e-10, stderr = 0.00136023603, R-squared = 0.326115, RMSE = 0.388700386316, Polynomial Regression: R-squared = 0.0927367, RMSE = 0.1818719990, Support Vector Regression: R-squared = 0.9680774158, RMSE = 0.0846001608. Figures 4 and 5 show different regressions for the first and second waves of Covid-19 in Kuwait.

**Fig. 4.**
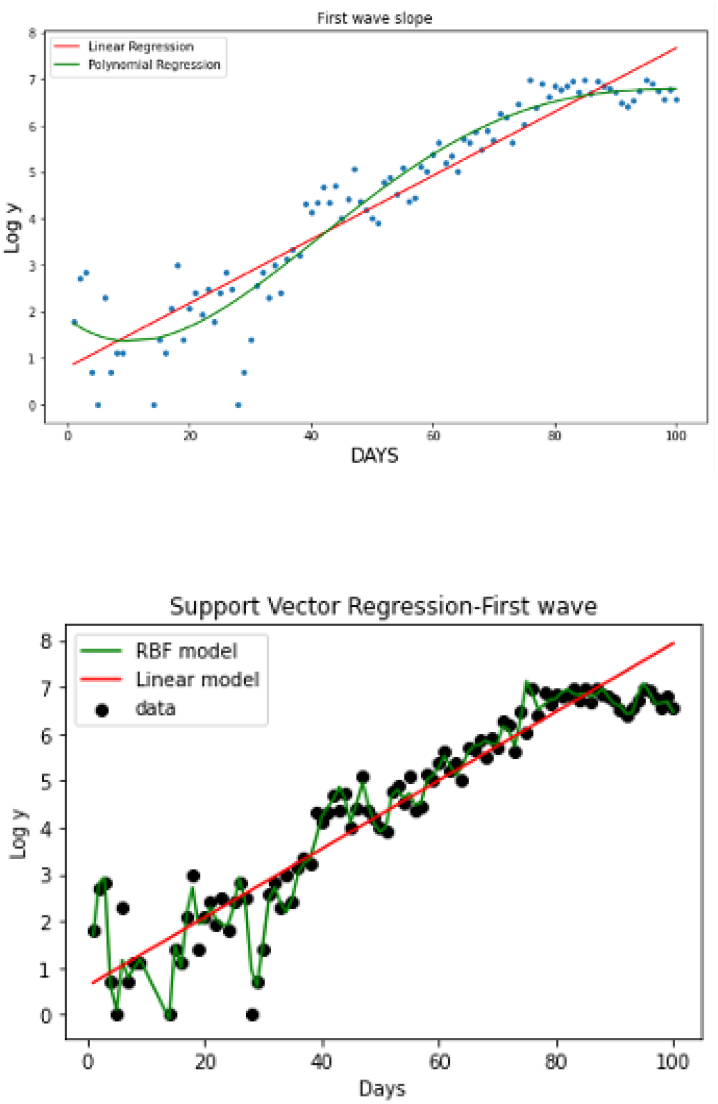
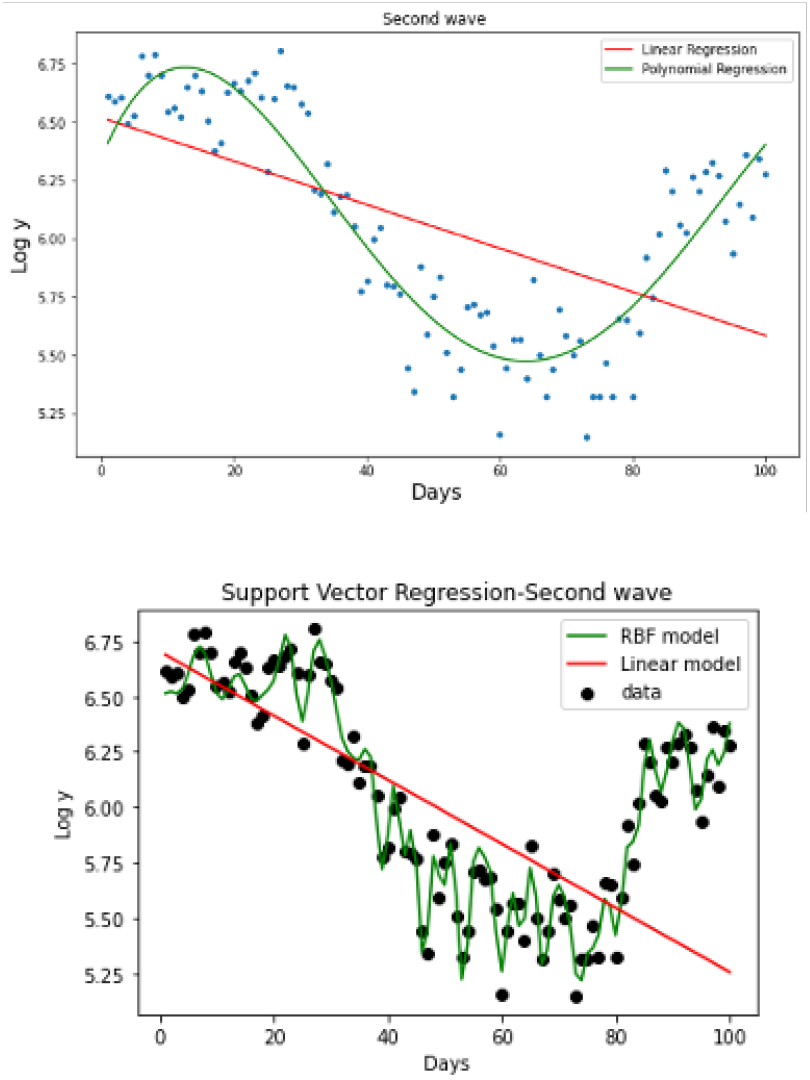
Log-linear, polynomial and support vector regressions for first wave start new cases and for second wave transition new cases. The origin of time corresponds to the corresponding wave start.

**Fig. 5.**
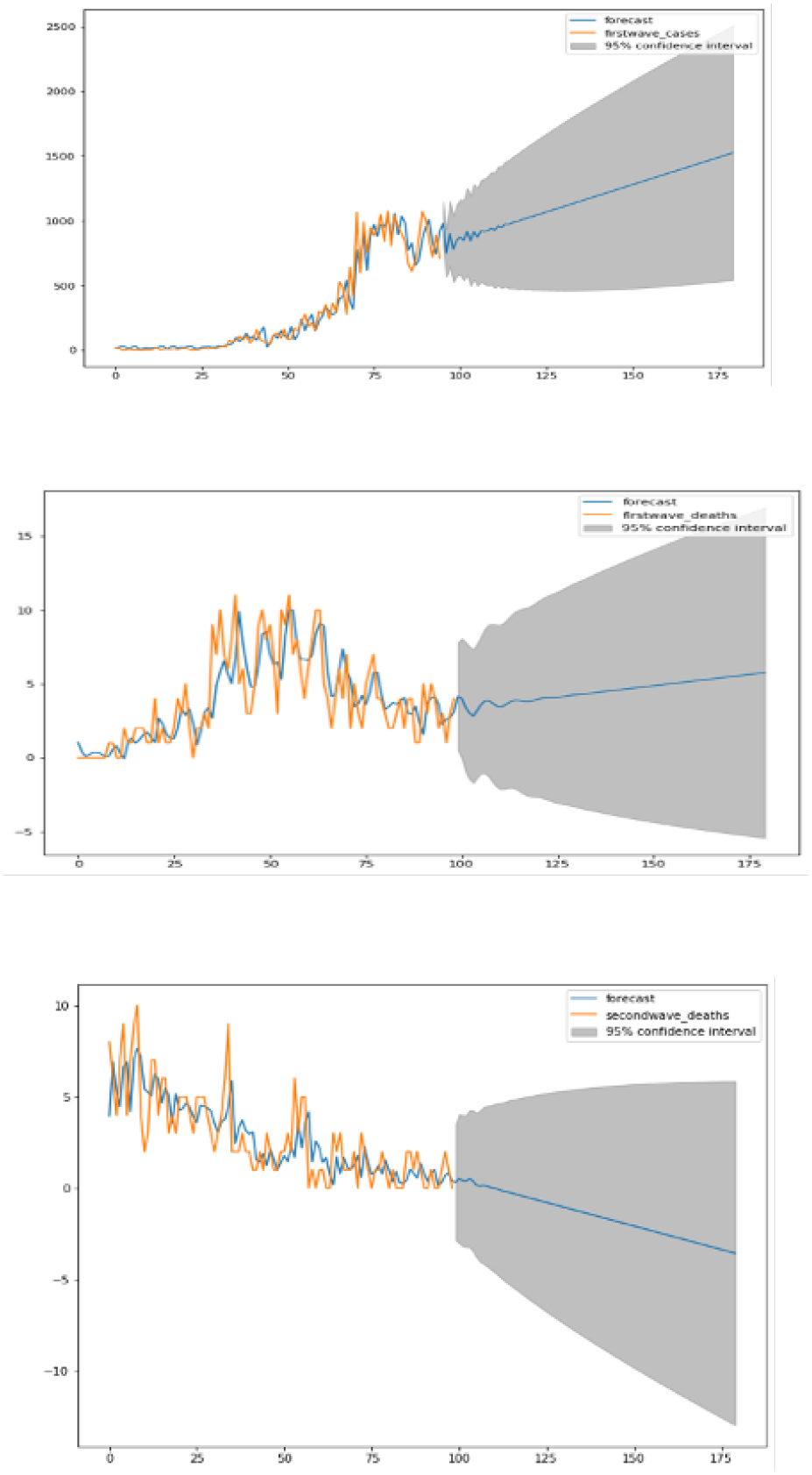
Up-ARIMA forecast for first wave start new cases, middle-ARIMA forecast for first wave deaths and bottom-ARIMA forecast for second wave deaths.

**Fig. 6.**
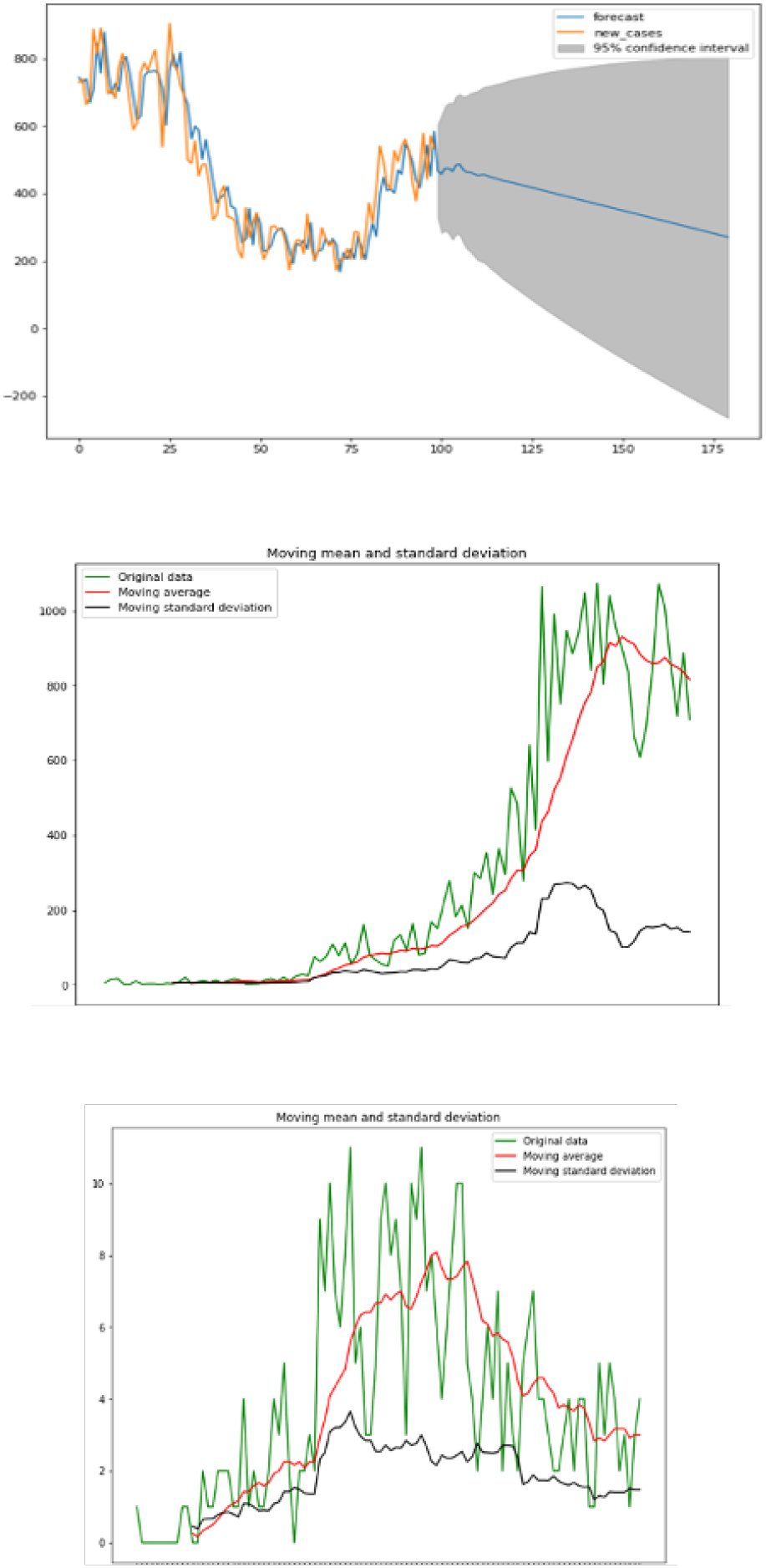
Up-ARIMA forecast for second wave transition new cases, Middle-Moving average and standard deviation for first wave start new cases and bottom-Moving average and standard deviation for first wave deaths. The origin of time corresponds to the corresponding wave start.

**Fig. 7.**
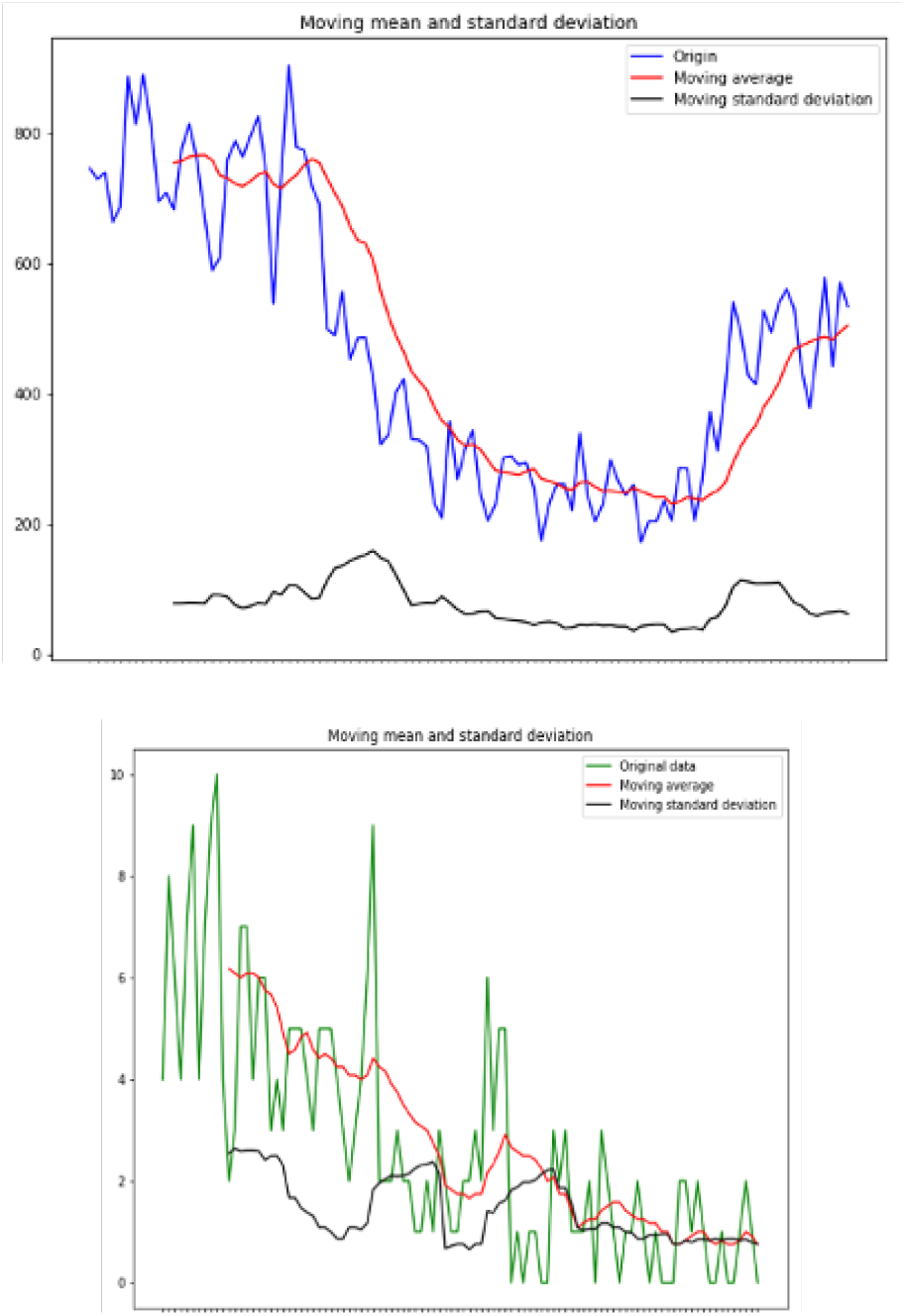
Up-Moving average and standard deviation for second wave transition new cases and down-Moving average and standard deviation for second wave deaths.The origin of time corresponds to the corresponding wave start.

The ARIMA initial autocorrelation slope is - 0.031 for the first wave start and -0.038 for the second wave transition, showing a stability of the autocorrelation at the start of these two phases.

### 3.2 Prophet® package Forecasting®

**Fig. 8.**
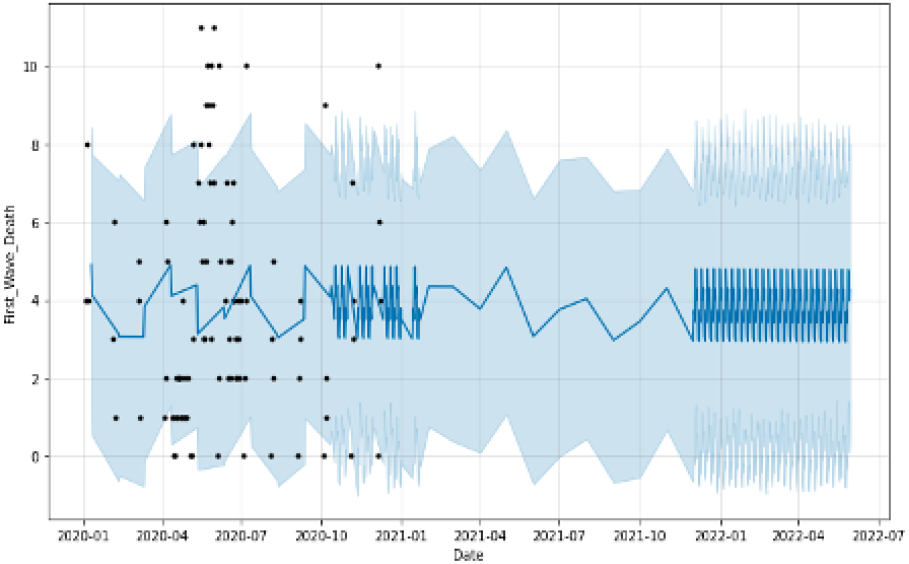
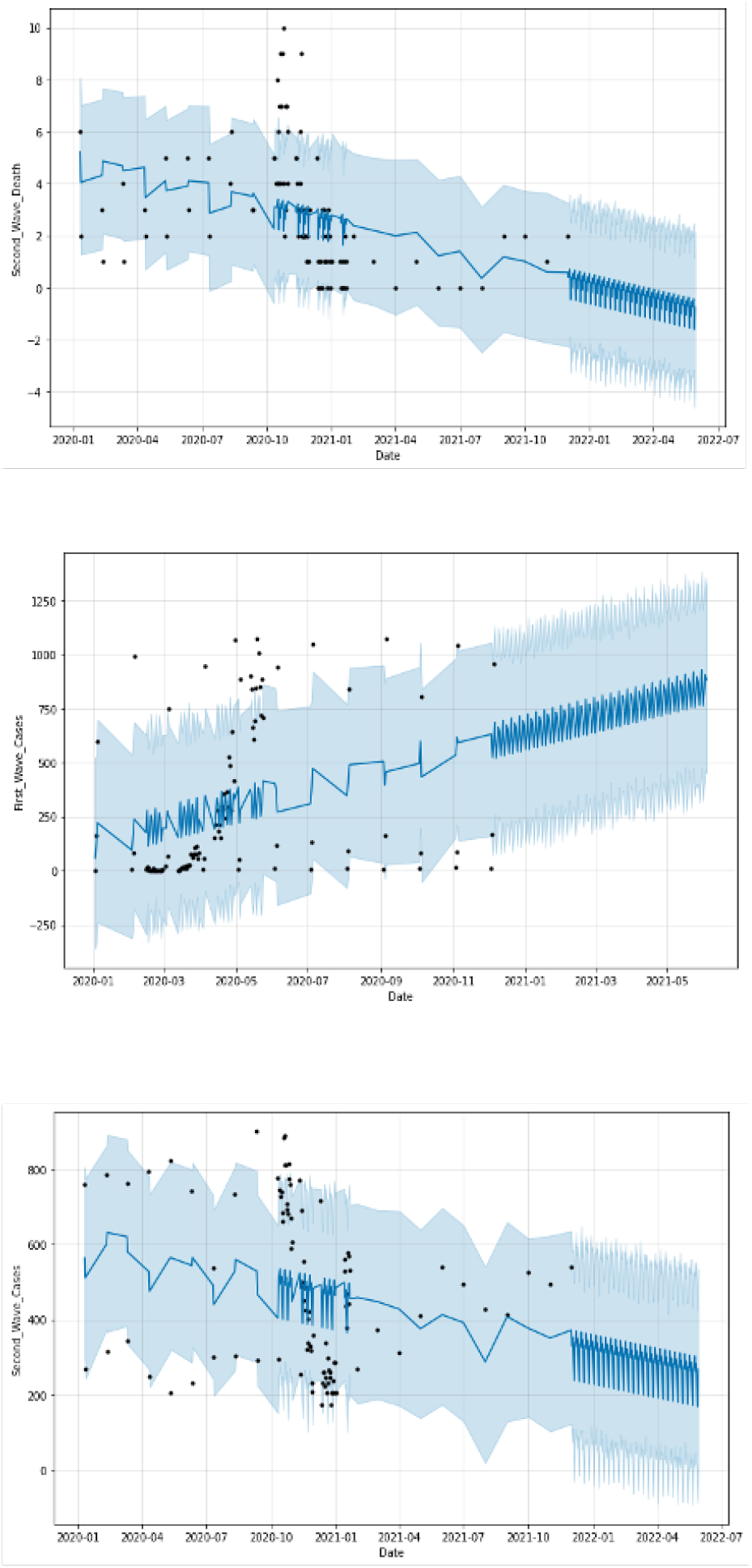
Prophet forecast for first wave start and second wave transition deaths and for first wave start and second wave transition new cases.

**Fig. 9.**
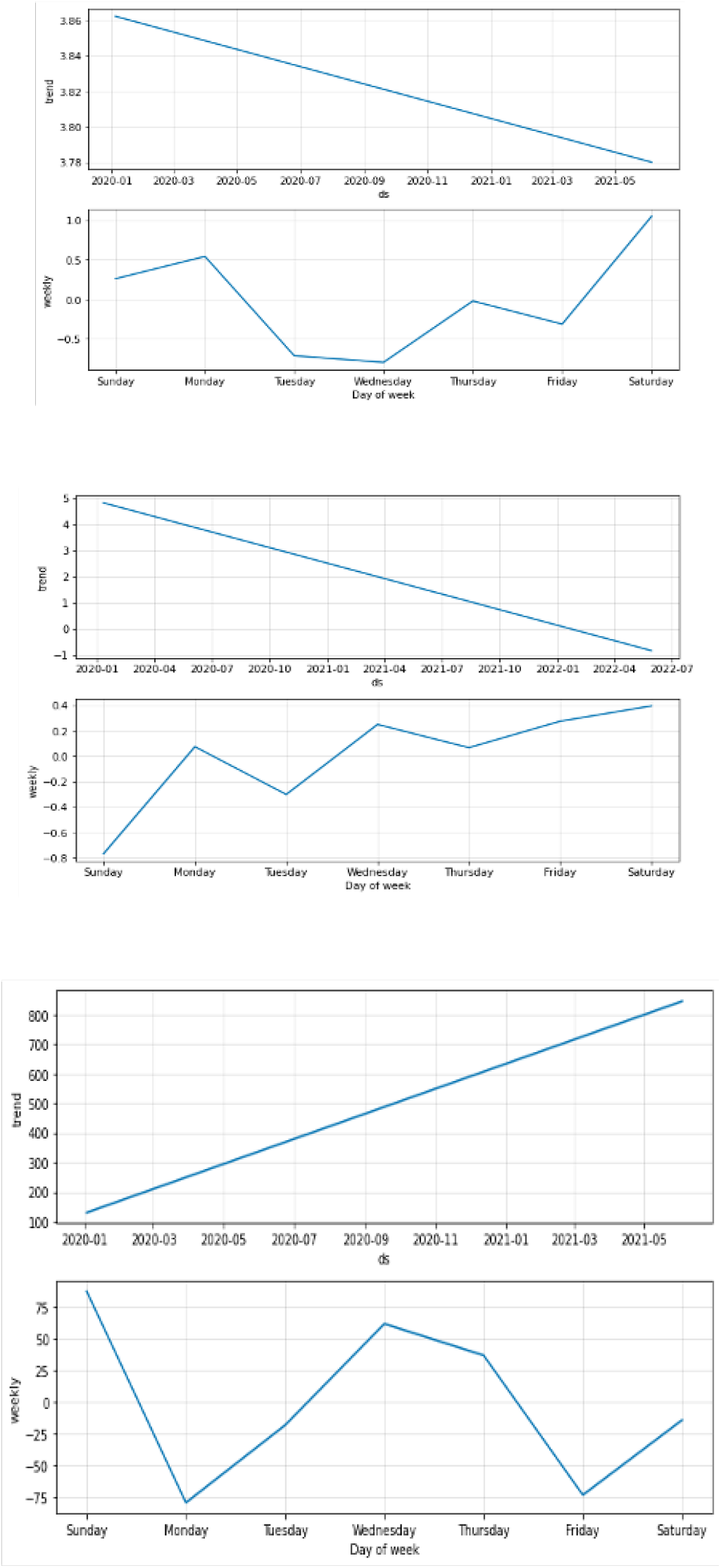
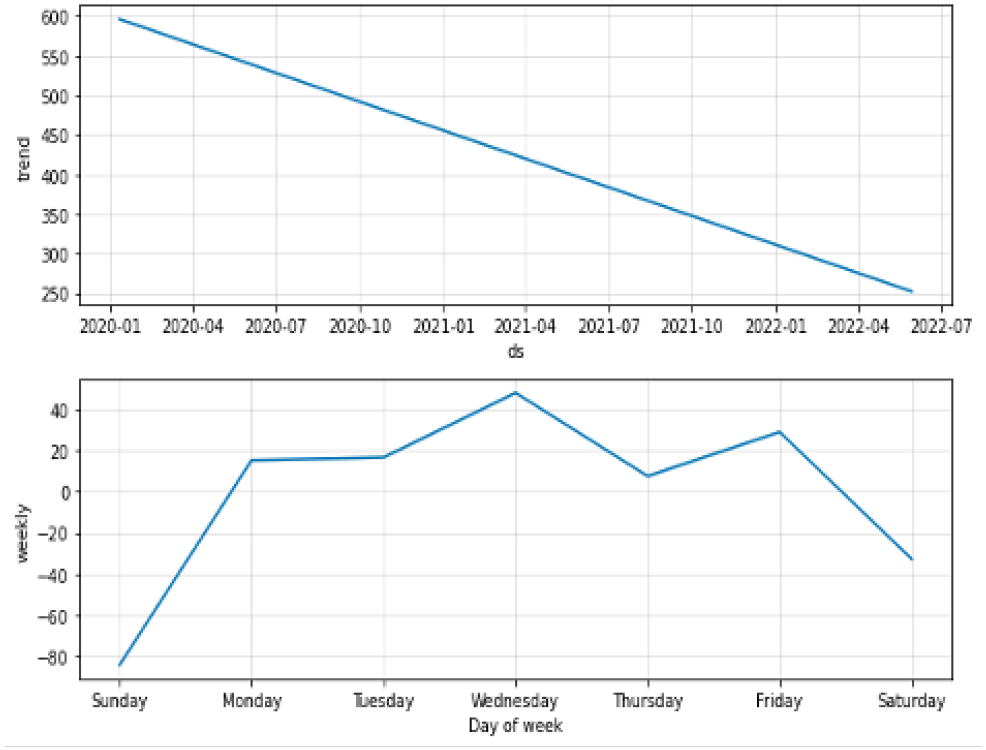
Trend plots for first wave start and second wave transition deaths, with stationary component (bottom); Same for first wave start and second wave transition new cases.

### 3.3 Exponential smoothing method

**Fig. 10.**
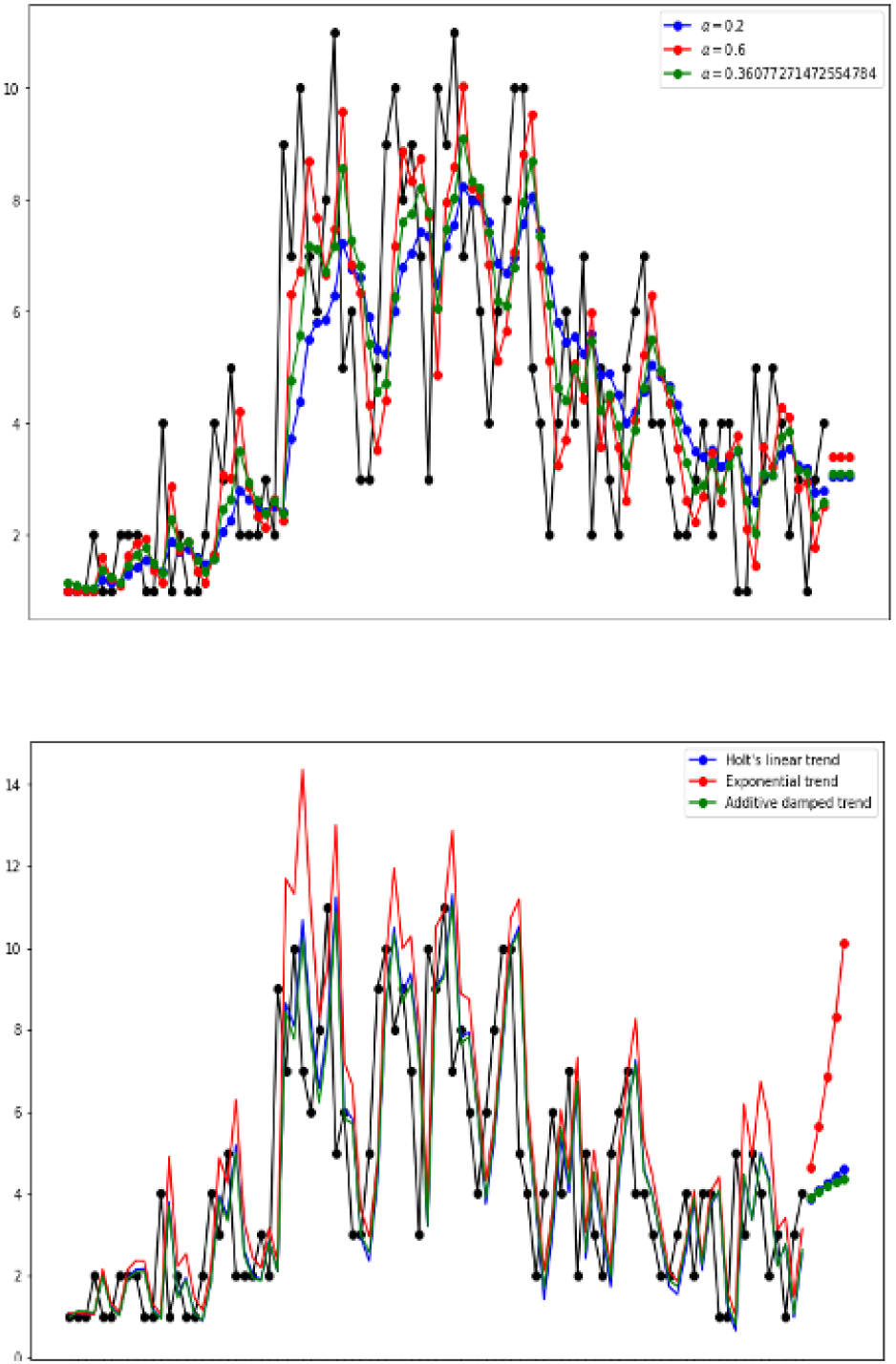
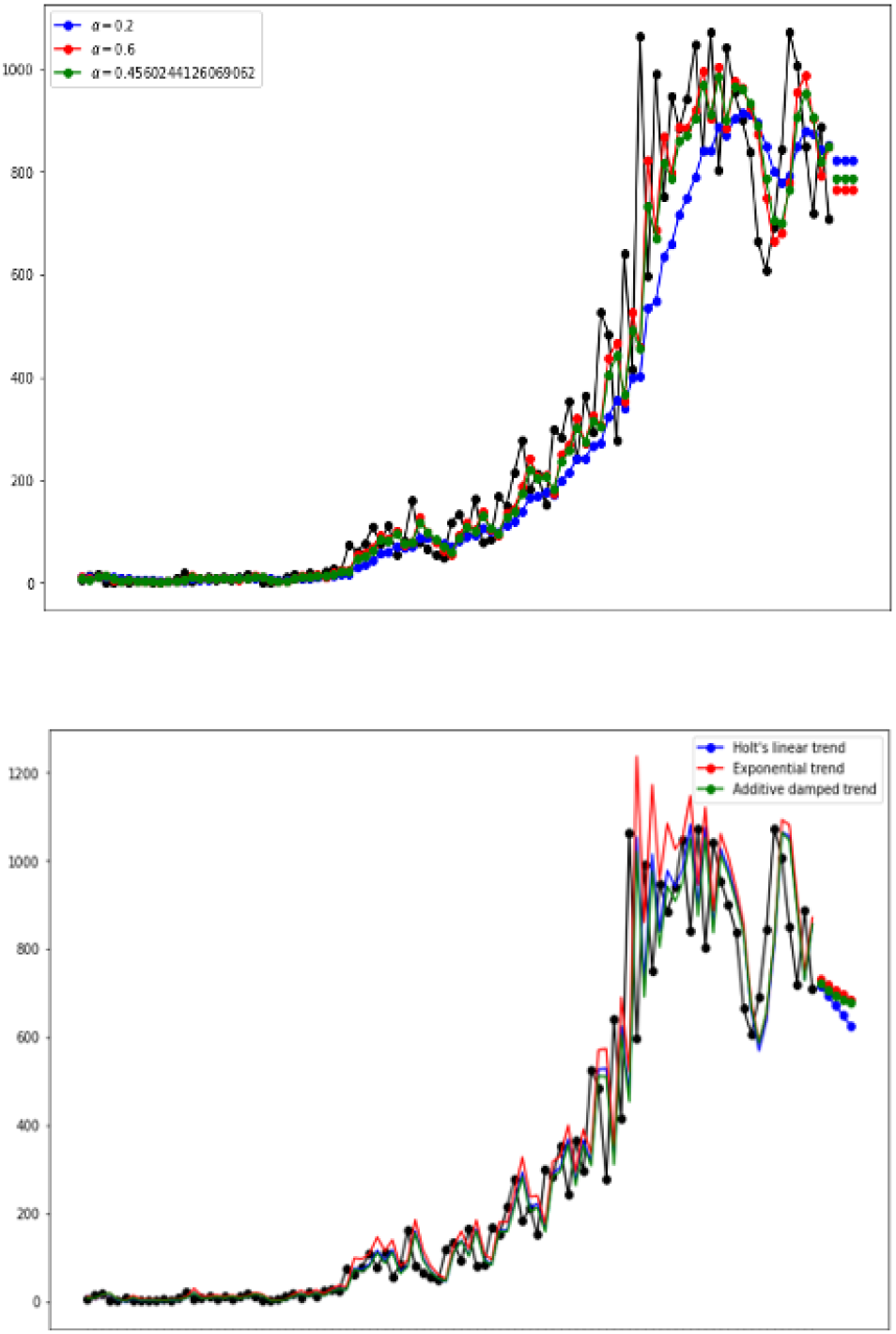
Exponential smoothing with different parameter values and comparison with other methods for first wave deaths and for first wave new cases. The origin of time corresponds to the corresponding wave start.

**Fig. 11.**
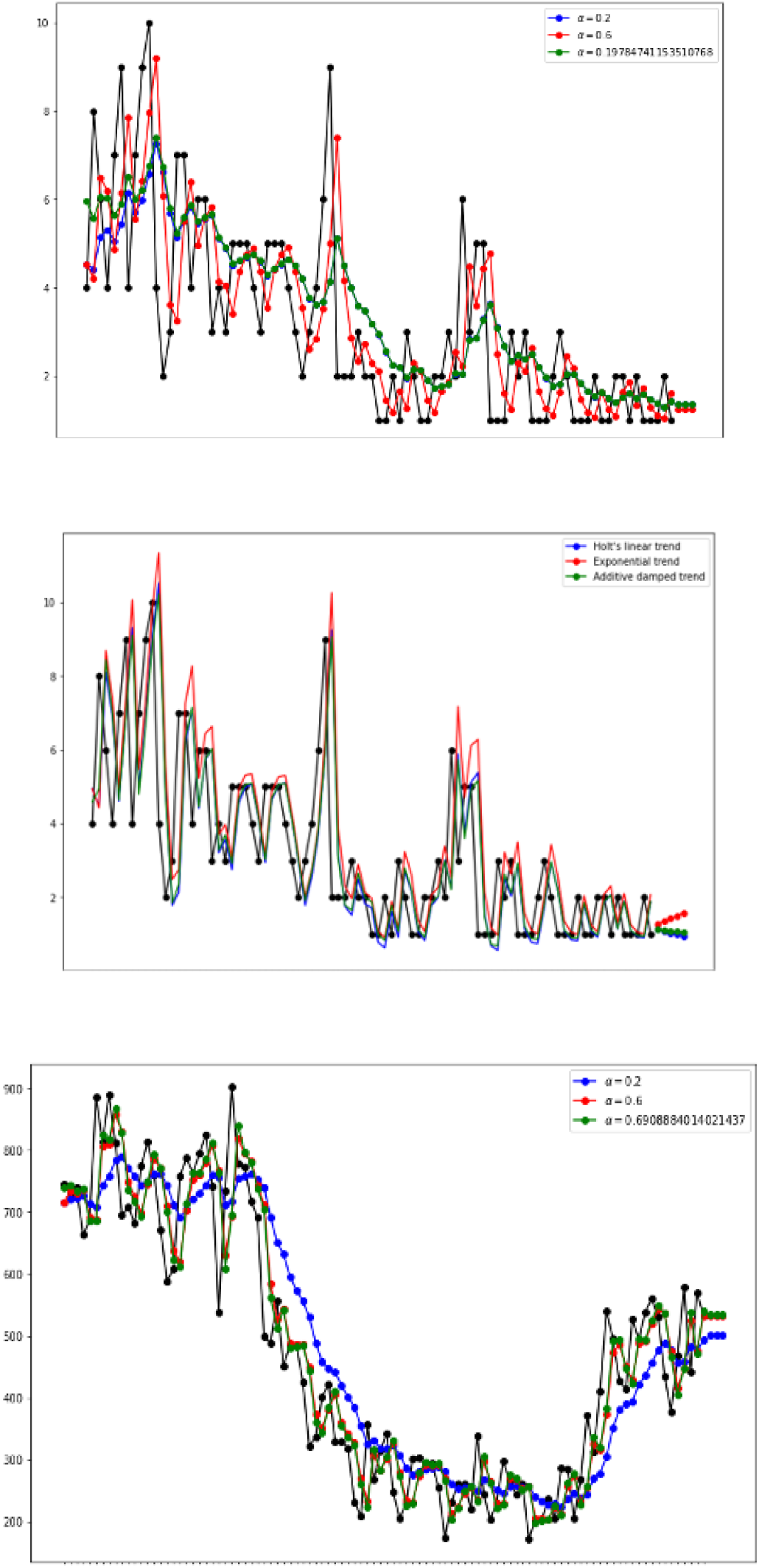
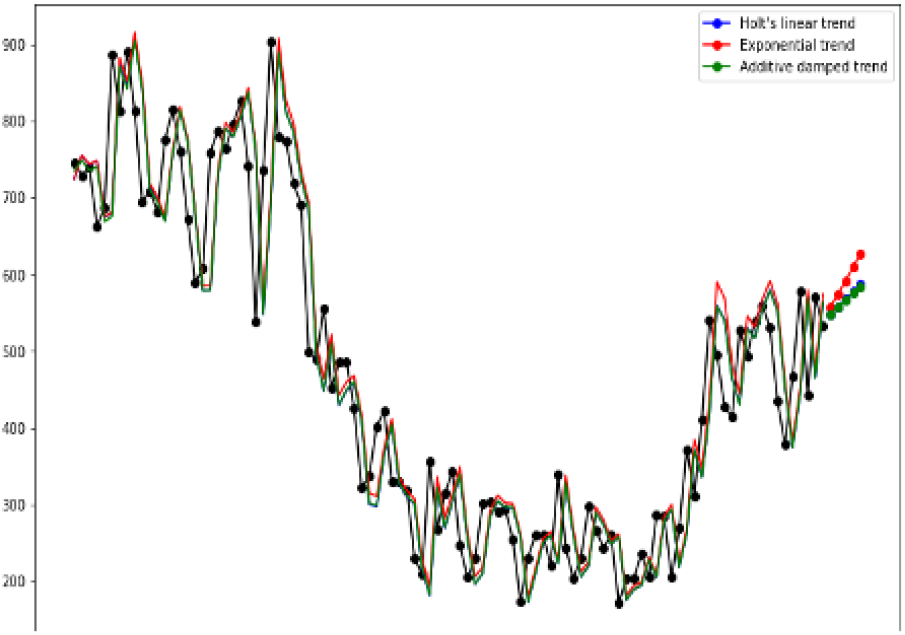
Exponential smoothing with different parameter values and comparison with other methods for second wave deaths and second wave new cases. The origin of time corresponds to the corresponding wave start.

#### 3.3.1 Holt’s method

**Fig. 12.**
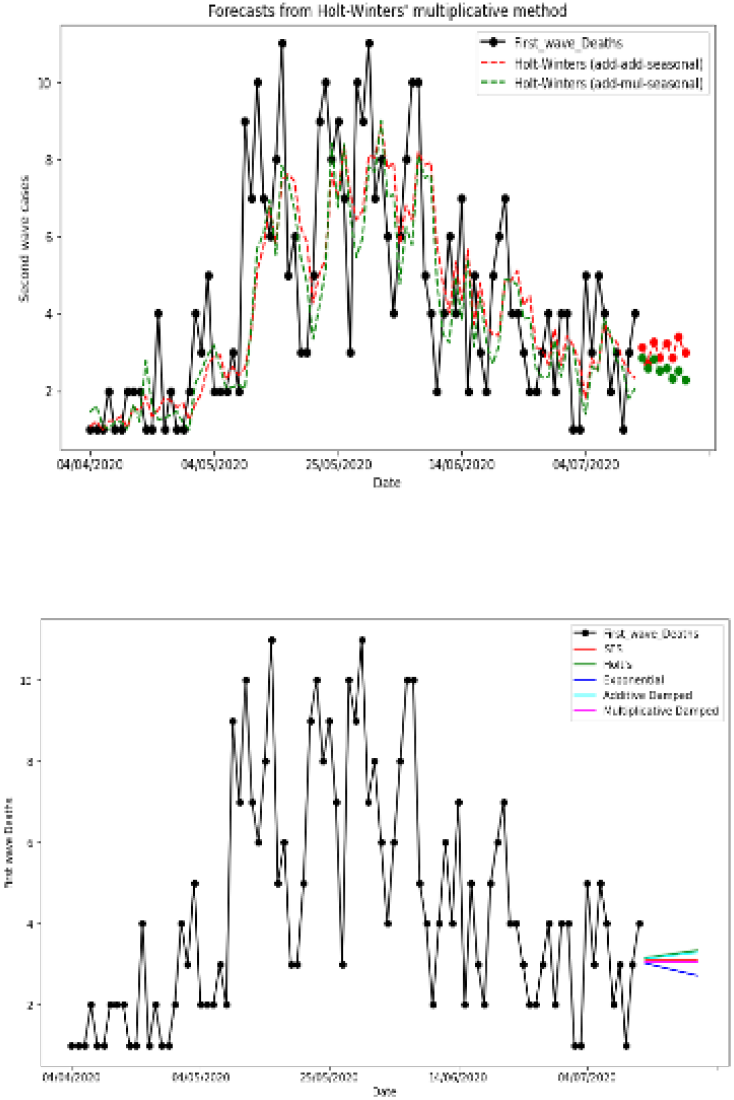
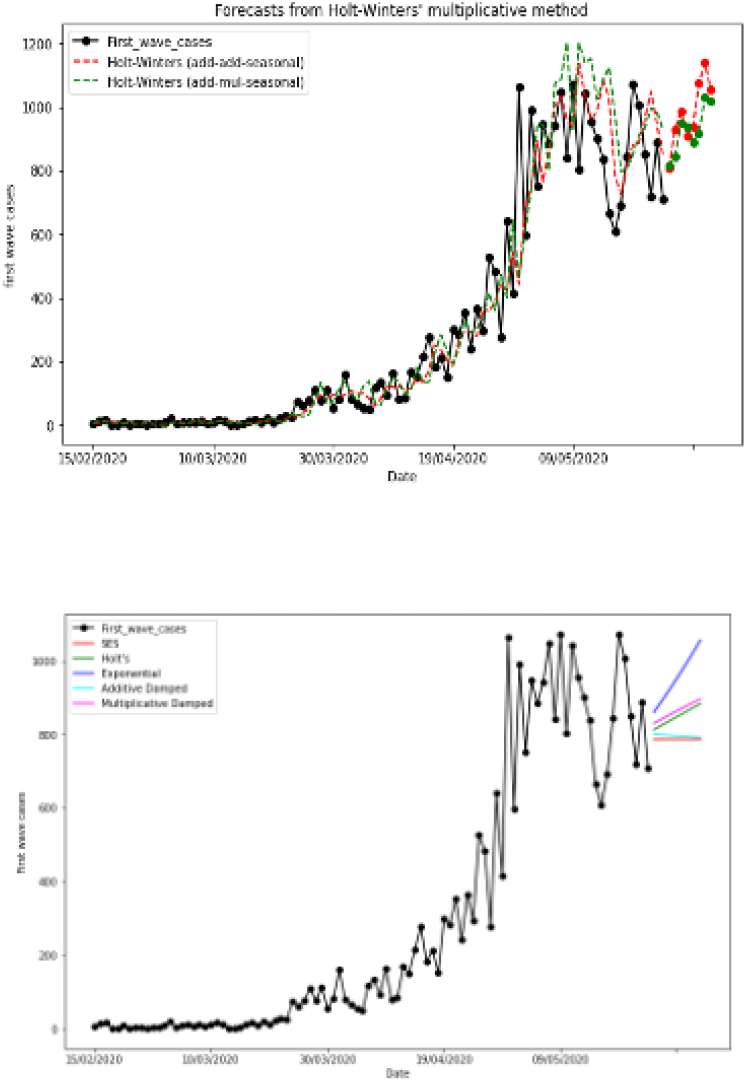
Holt’s method and comparison between different methods for first wave deaths and for first wave new cases.

**Fig. 13.**
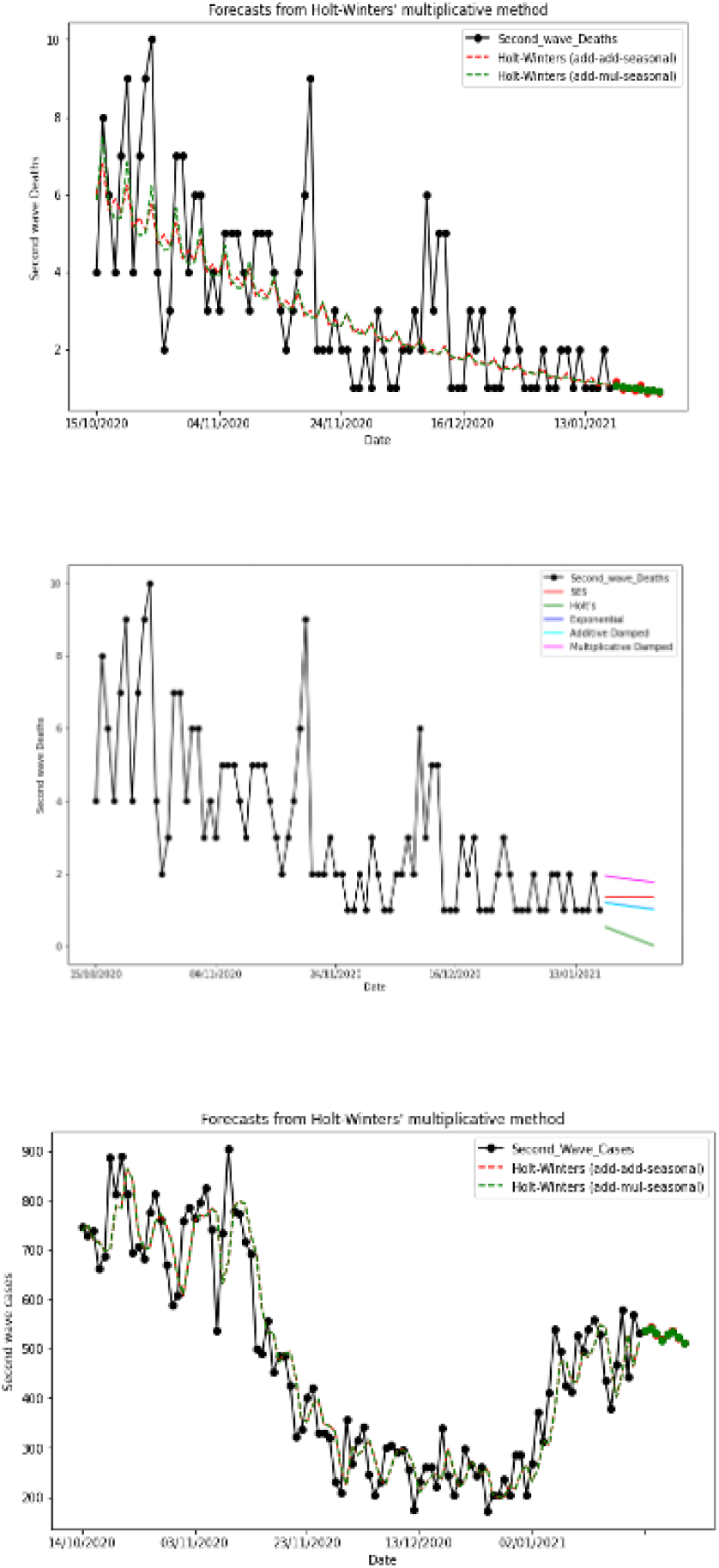
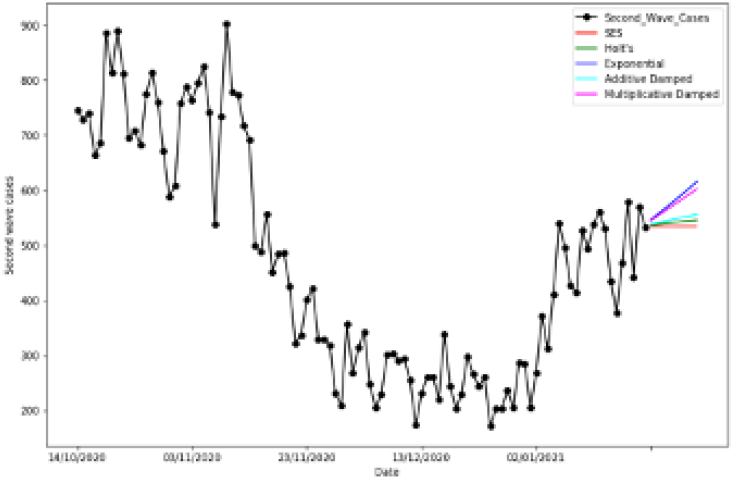
Holt’s method and comparison between different methods for second wave deaths and second wave new cases.

### 3.4 Neural Prophet method

**Fig. 14.**
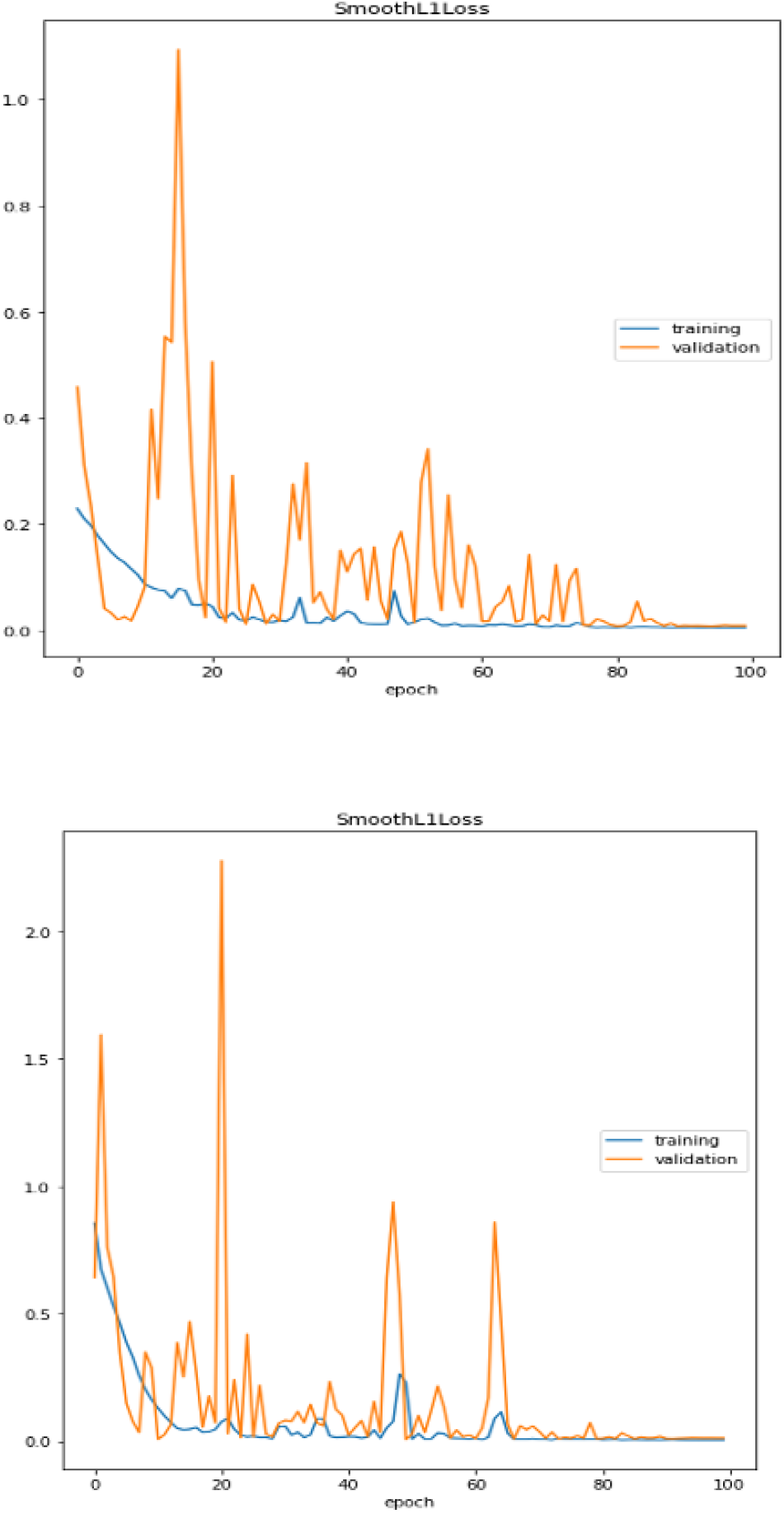
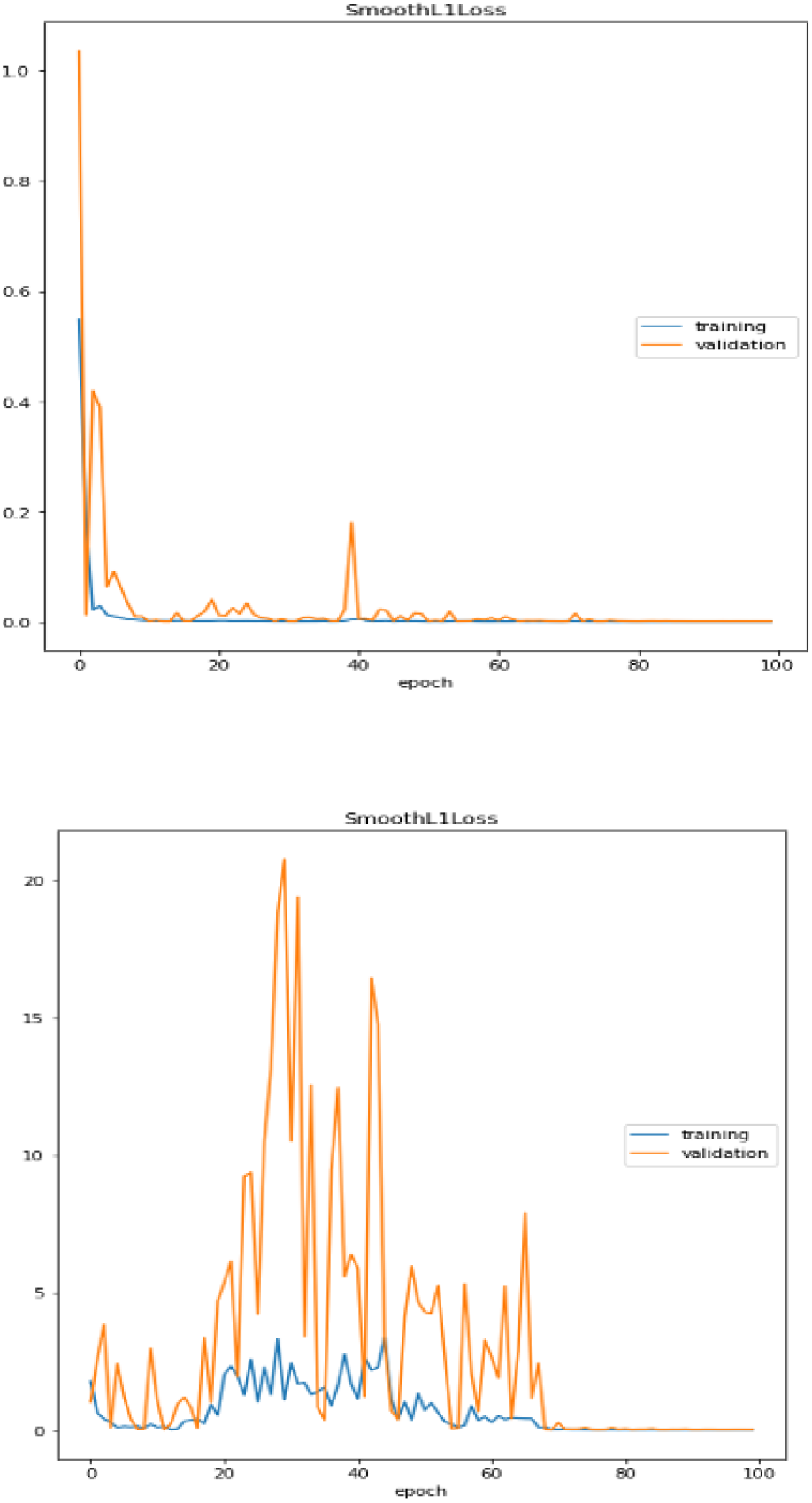
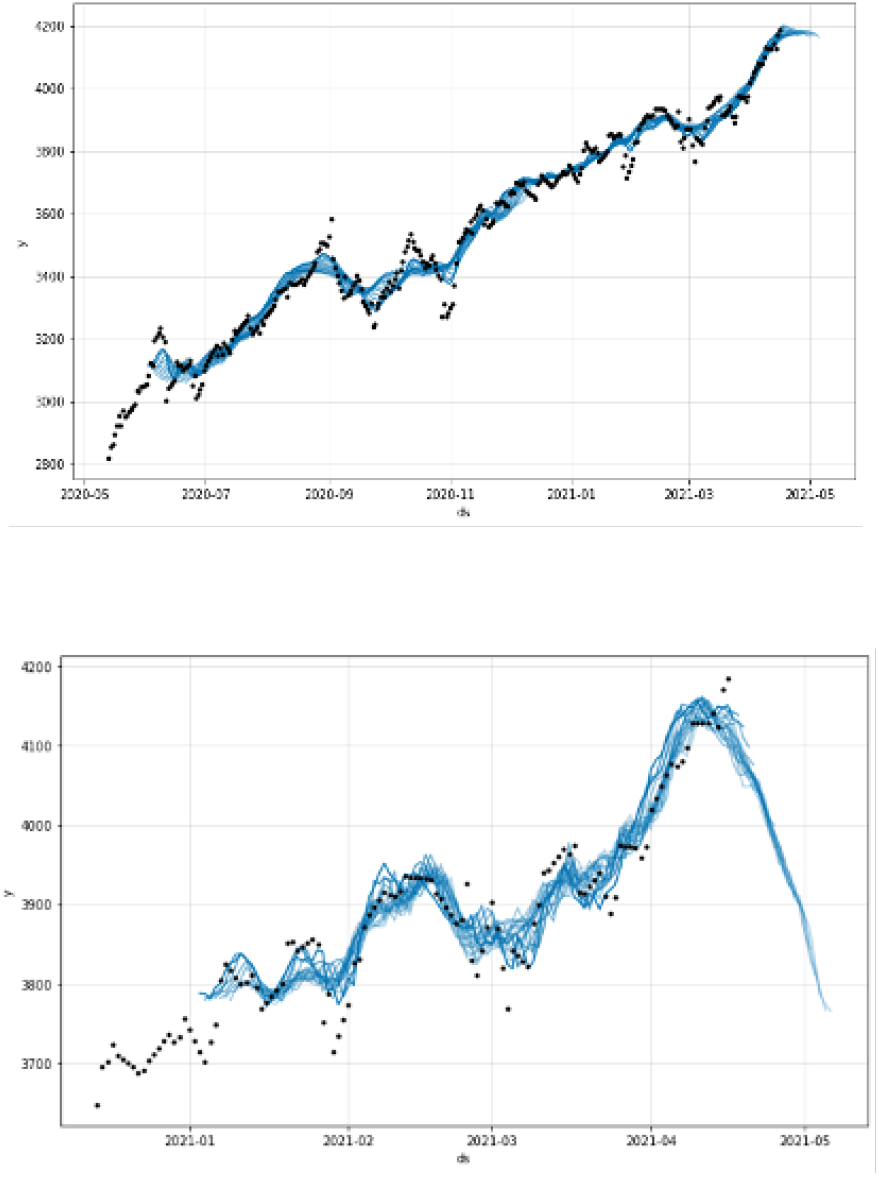
Loss plot for second wave deaths, second wave new cases, first wave deaths, first wave new cases and neural forecast for first and second wave cases.

## 4. Discussion and conclusion

The two phases of the Covid-19 outbreak in Kuwait present differences:

- the first wave start shows an increase of the daily new cases (with a peak of 1000 daily new cases) during about 60 days with a slope of the exponential regression equal to 0.07, followed by an endemic phase with a high mean number of daily new cases (about 600) and with a delay of about 15 days an increase of deaths,
- the second wave transition shows a decrease of the new cases during about 50 days with a slope of the exponential regression equal to -0.009, followed by an increase during about 20 days. But these two phases show a certain homogeneity in their stochastic structure at their beginning, because they have the same level for the variation coefficient (around 0.5), both for daily new cases number and for death number, and of autocorrelation initial slope (−0.031 for the first phase and -0.038 for the second).

We have been able to demonstrate different type of exponential smoothing models to give significance for recent observations and produce accurate forecasts while Holts model gives the trend and level of a time series and its computationally more efficient than double moving average, Winter holts model consider randomness using efficient smoothing process and its computationally efficient too.

We have also presented a loss plot for our neural network model and a loss plot helps to know the performance of the model. It was observed that our loss plot shows a good fit and converges during the weekly seasonality of the model. The plot of training loss decreases to a point of stability and also the plot of validation loss decreases to a point of stability and has a small gap (generalization gap) with the training loss.

## Data Availability

All data in the manuscript are comin from the public database Worldometer

## ACKNOLEDGEMENTS

The authors thank Pr. Pierre Magal and Dr. Hervé Seligmann for many fruitful discussions.

## Appendix

**Fig. 15.**
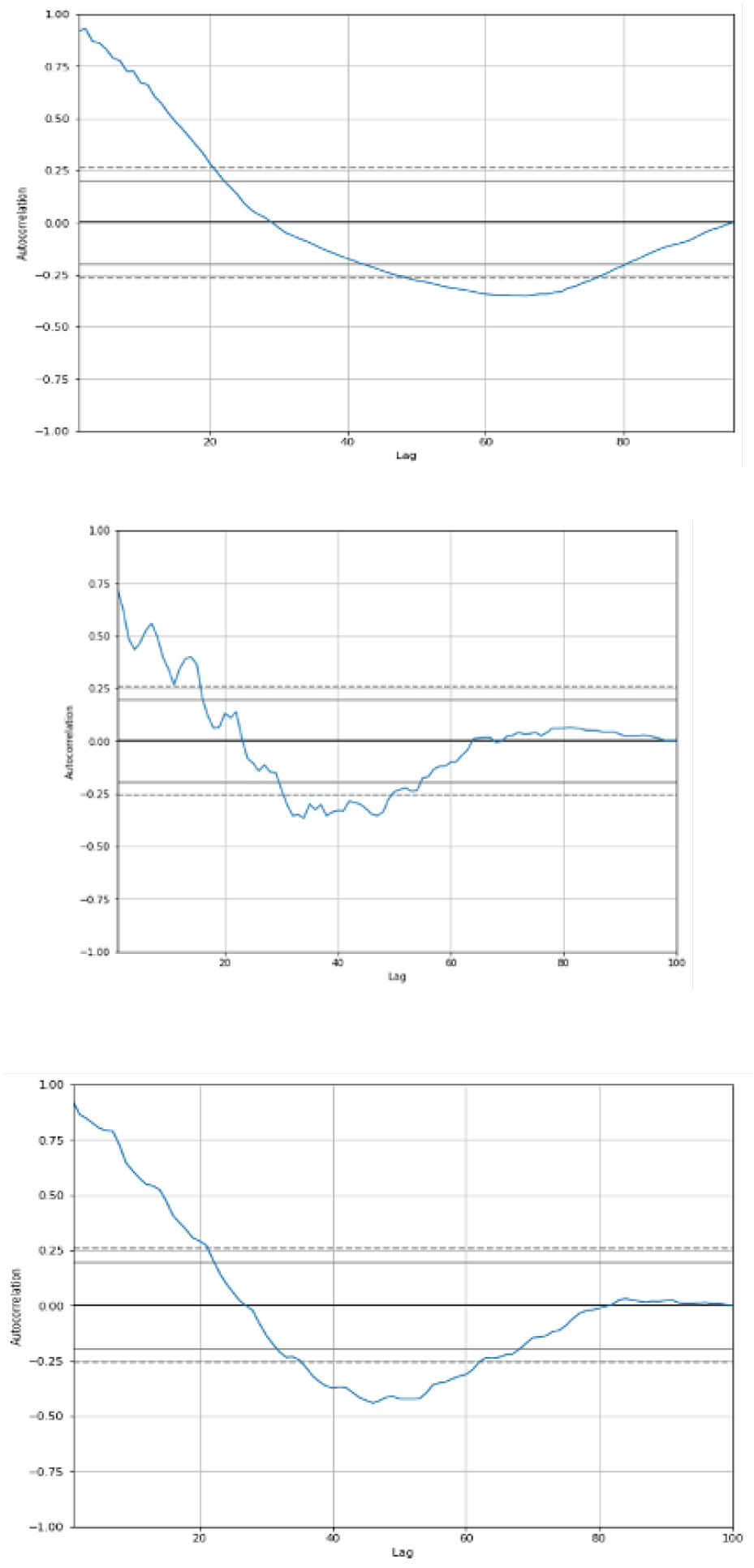
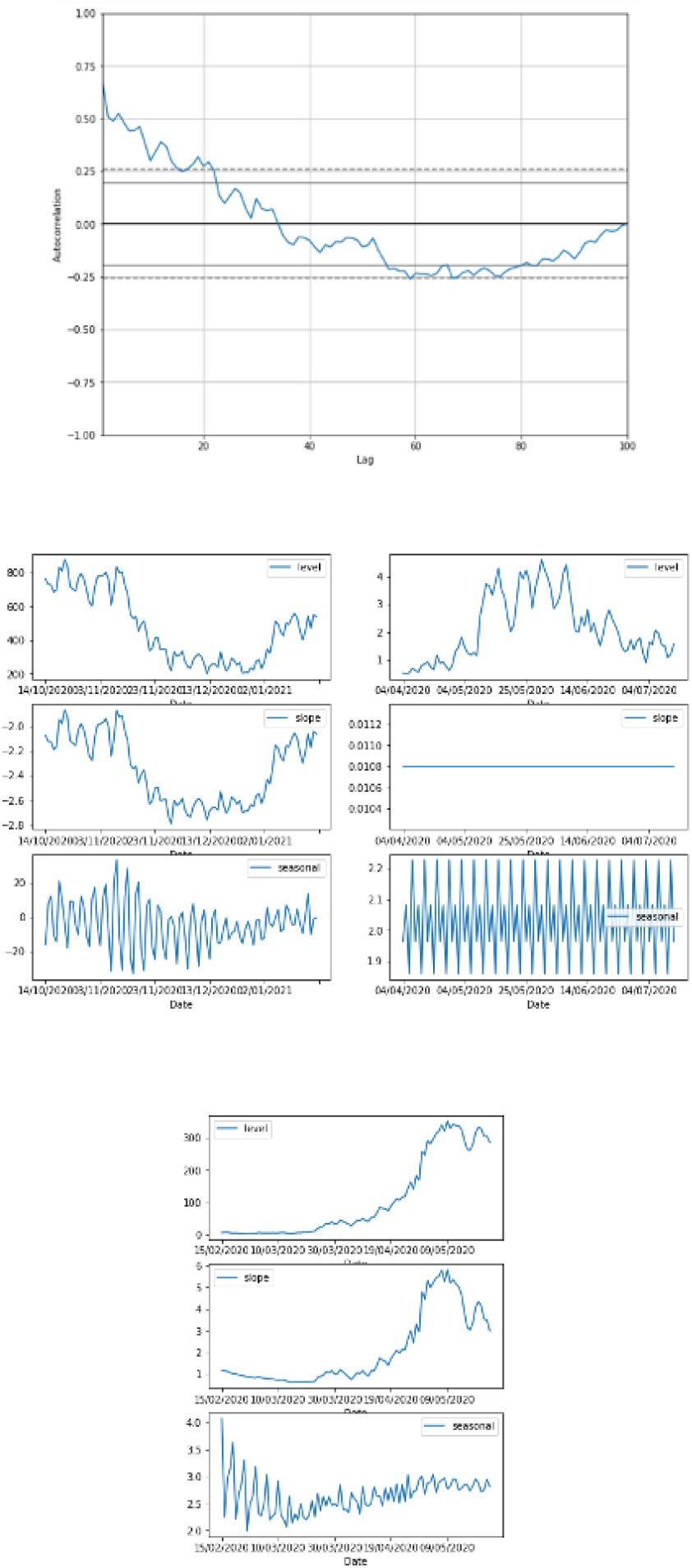
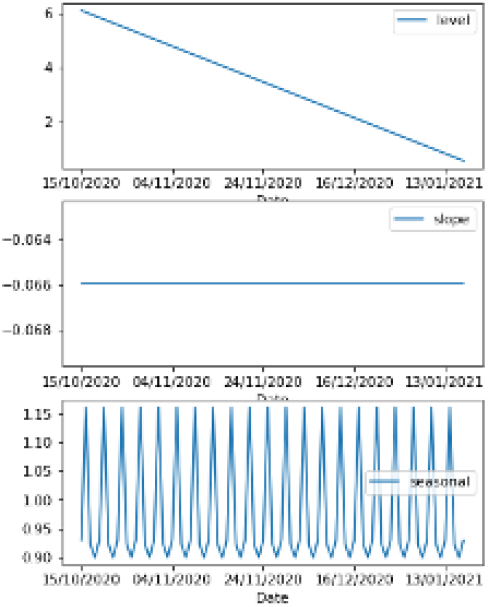
Autocorrelation curves for first wave start new cases and deaths, second wave transition new cases and deaths, level, slope and seasonal component for first wave new cases and for second wave new cases.

**Table 1.** Kuwait number of daily new cases predicted for October, 2020 (Note: PR means predicted range, OV means Observed value from worldometer and PV means predicted value).

| No | Date | PR | OV | PV | In range? |
| --- | --- | --- | --- | --- | --- |
| 1 | 11-10-20 | 150 ~ 651 | 548 | 404 | YES |
| 2 | 12-10-20 | 242 ~ 777 | 777 | 503 | YES |
| 3 | 14-10-20 | 276 ~ 789 | 532 | 535 | YES |
| 4 | 15-10-20 | 224 ~ 755 | 746 | 494 | YES |
| 5 | 16-10-20 | 249 ~ 769 | 729 | 515 | YES |
| 6 | 17-10-20 | 202 ~ 692 | 739 | 453 | NO |
| 7 | 18-10-20 | 161 ~ 657 | 663 | 401 | NO |
| 8 | 19-10-20 | 247 ~ 753 | 686 | 500 | YES |
| 9 | 20-10-20 | 240 ~ 777 | 886 | 501 | NO |
| 10 | 21-10-20 | 287 ~ 775 | 813 | 532 | NO |
| 11 | 22-10-20 | 228 ~ 738 | 889 | 491 | NO |
| 12 | 23-10-20 | 262 ~ 788 | 812 | 512 | NO |
| 13 | 24-10-20 | 210 ~ 703 | 695 | 450 | YES |
| 14 | 25-10-20 | 134 ~ 656 | 708 | 398 | NO |
| 15 | 26-10-20 | 244 ~ 754 | 682 | 498 | YES |
| 16 | 27-10-20 | 246 ~ 766 | 775 | 499 | YES |
| 17 | 28-10-20 | 270 ~ 792 | 814 | 530 | NO |
| 18 | 29-10-20 | 232 ~ 754 | 760 | 489 | NO |
| 19 | 30-10-20 | 248 ~ 766 | 671 | 510 | YES |
| 20 | 31-10-20 | 183 ~ 728 | 589 | 447 | YES |

**Table 2.**
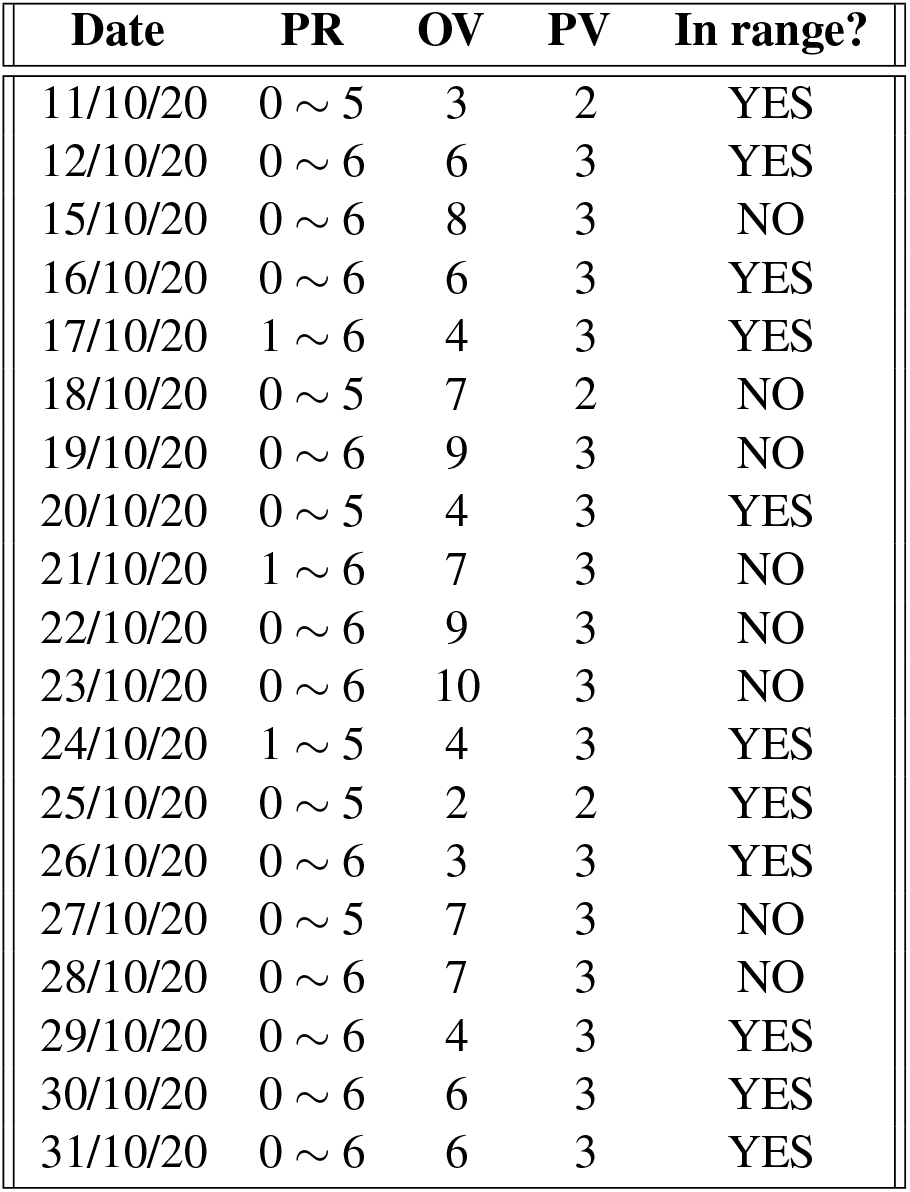
Kuwait number of daily deaths predicted for October, 2020 (Note: PR means predicted range, OV means Observed value from worldometer and PV means predicted value).

| Date | PR | OV | PV | In range? |
| --- | --- | --- | --- | --- |
| 11/10/20 | 0 ~ 5 | 3 | 2 | YES |
| 12/10/20 | 0 ~ 6 | 6 | 3 | YES |
| 15/10/20 | 0 ~ 6 | 8 | 3 | NO |
| 16/10/20 | 0 ~ 6 | 6 | 3 | YES |
| 17/10/20 | 1 ~ 6 | 4 | 3 | YES |
| 18/10/20 | 0 ~ 5 | 7 | 2 | NO |
| 19/10/20 | 0 ~ 6 | 9 | 3 | NO |
| 20/10/20 | 0 ~ 5 | 4 | 3 | YES |
| 21/10/20 | 1 ~ 6 | 7 | 3 | NO |
| 22/10/20 | 0 ~ 6 | 9 | 3 | NO |
| 23/10/20 | 0 ~ 6 | 10 | 3 | NO |
| 24/10/20 | 1 ~ 5 | 4 | 3 | YES |
| 25/10/20 | 0 ~ 5 | 2 | 2 | YES |
| 26/10/20 | 0 ~ 6 | 3 | 3 | YES |
| 27/10/20 | 0 ~ 5 | 7 | 3 | NO |
| 28/10/20 | 0 ~ 6 | 7 | 3 | NO |
| 29/10/20 | 0 ~ 6 | 4 | 3 | YES |
| 30/10/20 | 0 ~ 6 | 6 | 3 | YES |
| 31/10/20 | 0 ~ 6 | 6 | 3 | YES |

**Table 3.** Kuwait number of daily deaths and daily cases predicted for April and May, 2021 (Note: PRD means predicted range for daily deaths, PVD means predicted values for daily deaths, PRC predicted range for daily cases and PVC means predicted value for daily cases).

| No | Date | PRD | PVD | PRC | PVC |
| --- | --- | --- | --- | --- | --- |
| 1 | 17-04-21 | 1 ~ 8 | 5 | 344 ~ 1223 | 766 |
| 2 | 18-04-21 | 1 ~ 8 | 4 | 467 ~ 1303 | 869 |
| 3 | 19-04-21 | 1 ~ 8 | 3 | 267 ~ 1164 | 704 |
| 4 | 22-04-21 | 0 ~ 7 | 4 | 380 ~ 1276 | 824 |
| 5 | 23-04-21 | 0 ~ 7 | 3 | 237 ~ 1172 | 716 |
| 6 | 24-04-21 | 1 ~ 9 | 5 | 345 ~ 1241 | 776 |
| 7 | 25-04-21 | 0 ~ 8 | 4 | 445 ~ 1303 | 879 |
| 8 | 26-04-21 | 1 ~ 8 | 4 | 268 ~ 1131 | 714 |
| 9 | 27-04-21 | 0 ~ 7 | 3 | 336 ~ 1233 | 776 |
| 10 | 28-04-21 | 0 ~ 7 | 3 | 397 ~ 1280 | 857 |
| 11 | 29-04-21 | 0 ~ 8 | 4 | 395 ~ 1271 | 834 |
| 12 | 30-04-21 | 0 ~ 7 | 3 | 281 ~ 1166 | 725 |
| 13 | 01-05-21 | 1 ~ 8 | 5 | 340 ~ 1216 | 786 |
| 14 | 02-05-21 | 0 ~ 8 | 4 | 439 ~ 1311 | 888 |
| 15 | 03-05-21 | 0 ~ 8 | 4 | 282 ~ 1151 | 723 |
| 16 | 04-05-21 | 0 ~ 7 | 3 | 369 ~ 1228 | 786 |
| 17 | 05-05-21 | 0 ~ 7 | 3 | 455 ~ 1313 | 867 |
| 18 | 06-05-21 | 0 ~ 8 | 4 | 387 ~ 1274 | 844 |
| 19 | 07-05-21 | 0 ~ 7 | 3 | 280 ~ 1178 | 735 |
| 20 | 08-05-21 | 1 ~ 8 | 5 | 357 ~ 1215 | 795 |
| 21 | 09-05-21 | 0 ~ 8 | 4 | 478 ~ 1342 | 898 |
| 22 | 10-05-21 | 1 ~ 8 | 4 | 291 ~ 1155 | 733 |
| 23 | 11-05-21 | 0 ~ 7 | 3 | 371 ~ 1230 | 795 |
| 24 | 12-05-21 | 0 ~ 6 | 3 | 457 ~ 1345 | 877 |
| 25 | 13-05-21 | 0 ~ 8 | 4 | 447 ~ 1302 | 853 |
| 26 | 14-05-21 | 0 ~ 7 | 3 | 330 ~ 1211 | 744 |
| 27 | 15-05-21 | 1 ~ 9 | 5 | 383 ~ 1259 | 805 |
| 28 | 16-05-21 | 0 ~ 8 | 4 | 448 ~ 1369 | 908 |
| 29 | 17-05-21 | 1 ~ 8 | 4 | 308 ~ 1193 | 743 |
| 30 | 18-05-21 | 0 ~ 7 | 3 | 381 ~ 1245 | 805 |
| 31 | 19-05-21 | 0 ~ 7 | 3 | 450 ~ 1333 | 886 |
| 32 | 20-05-21 | 0 ~ 7 | 4 | 441 ~ 1291 | 863 |
| 33 | 21-05-21 | 0 ~ 7 | 3 | 299 ~ 1184 | 754 |
| 34 | 22-05-21 | 1 ~ 9 | 5 | 332 ~ 1284 | 815 |
| 35 | 23-05-21 | 0 ~ 7 | 4 | 467 ~ 1354 | 917 |
| 36 | 24-05-21 | 1 ~ 8 | 4 | 279 ~ 1184 | 753 |
| 37 | 25-05-21 | 0 ~ 7 | 3 | 362 ~ 1262 | 815 |
| 38 | 26-05-21 | 0 ~ 7 | 3 | 494 ~ 1332 | 896 |
| 39 | 27-05-21 | 0 ~ 7 | 8 | 406 ~ 1299 | 873 |
| 40 | 28-05-21 | 0 ~ 7 | 3 | 340 ~ 1213 | 764 |
| 41 | 29-05-21 | 1 ~ 9 | 5 | 367 ~ 1241 | 824 |
| 42 | 30-10-20 | 0 ~ 7 | 4 | 510 ~ 1375 | 927 |
| 43 | 31-10-20 | 1 ~ 8 | 4 | 328 ~ 1207 | 762 |

---

**Table 4.** Comparison of two methods for daily cases and deaths (Note: ESC means exponential smoothing model predicted value for daily cases, OVC means Observed value for daily cases from worldometer, PMC means prophet model predicted value for daily cases, ESD means exponential smoothing model predicted value for daily deaths, OVD means Observed value for daily deaths from worldometer, PMD means prophet model predicted value for daily deaths and MSE is mean square error).

| Date | ESC | OVC | PMC | OVD | ESD | PMD |
| --- | --- | --- | --- | --- | --- | --- |
| 15-10-20 | 740 | 746 | 494 | 8 | 6 | 3 |
| 16-10-20 | 739 | 729 | 515 | 6 | 6 | 3 |
| 17-10-20 | 718 | 739 | 453 | 4 | 6 | 3 |
| 18-10-20 | 665 | 663 | 401 | 7 | 6 | 1 |
| 19-10-20 | 702 | 686 | 500 | 9 | 6 | 3 |
| 20-10-20 | 841 | 886 | 501 | 4 | 5 | 3 |
| 21-10-20 | 795 | 813 | 532 | 7 | 5 | 3 |
| 22-10-20 | 859 | 889 | 491 | 9 | 5 | 3 |
| 23-10-20 | 858 | 812 | 512 | 10 | 6 | 3 |
| 24-10-20 | 724 | 695 | 450 | 4 | 5 | 3 |
| 25-10-20 | 696 | 708 | 398 | 2 | 5 | 2 |
| 26-10-20 | 669 | 682 | 498 | 3 | 5 | 3 |
| 27-10-20 | 773 | 775 | 499 | 7 | 6 | 3 |
| 28-10-20 | 801 | 814 | 530 | 7 | 5 | 3 |
| 29-10-20 | 755 | 760 | 489 | 4 | 5 | 3 |
| 30-10-20 | 685 | 671 | 510 | 6 | 5 | 3 |
| 31-10-20 | 634 | 589 | 447 | 6 | 5 | 3 |
| MSE | 575.0 |  | 72721.5 |  | 14.7 | 4.5 |

